# Increasing coordination and responsivity of emotion-related brain regions with a heart rate variability biofeedback randomized trial

**DOI:** 10.1101/2021.09.28.21264206

**Authors:** Kaoru Nashiro, Jungwon Min, Hyun Joo Yoo, Christine Cho, Shelby L. Bachman, Shubir Dutt, Julian F. Thayer, Paul Lehrer, Tiantian Feng, Noah Mercer, Padideh Nasseri, Diana Wang, Catie Chang, Vasilis Z. Marmarelis, Shri Narayanan, Daniel A. Nation, Mara Mather

## Abstract

Heart rate variability is a robust biomarker of emotional well-being, consistent with the shared brain networks regulating emotion regulation and heart rate. While high heart rate oscillatory activity clearly indicates healthy regulatory brain systems, can increasing this oscillatory activity also affect brain function? To test this possibility, we randomly assigned 106 young adult participants to one of two 5-week interventions involving daily biofeedback that either increased heart rate oscillations (Osc+ condition) or had little effect on heart rate oscillations (Osc- condition) and examined effects on brain activity during rest and during regulating emotion. While there were no significant changes in the right amygdala-medial prefrontal cortex (MPFC) functional connectivity (our primary outcome), the Osc+ intervention increased left amygdala-MPFC functional connectivity and functional connectivity in emotion-related resting-state networks during rest. It also increased down-regulation of activity in somatosensory brain regions during an emotion regulation task. The Osc- intervention did not have these effects. In this healthy cohort, the two conditions did not differentially affect anxiety, depression or mood. These findings indicate that heart rate oscillatory activity not only provides a measure of the current state of regulatory brain systems but also changes emotion network coordination in the brain.

**Significance Statement:** People whose breathing makes their heart rate oscillate more (leading to higher heart rate variability or HRV) generally have better regulated emotion. Thus, HRV may indicate functioning of brain networks regulating emotion and internal body states. But heart rate oscillations may not only reflect brain regulatory networks but also help shape these networks. We randomly assigned participants to practice either increasing heart rate oscillations using slow-paced breathing or decreasing them using personalized strategies. Daily practice increasing heart rate oscillations affected brain activity in emotion-related networks even during times when participants breathed no differently than the comparison group. Thus, HRV is more than just an outcome measure--it can help shape the subsequent functioning of emotion-related brain networks.

---

Hundreds of previous studies have identified vagal HRV at rest as one of the best indicators of well-being (1-3). Vagal HRV refers to HRV measures that reflect relatively high frequency (HF) heart rate oscillations (HF-HRV) or changes in the length of adjacent intervals between heart beats (root mean square of successive differences; RMSSD). These relatively faster changes in heart rate are transmitted by the vagus nerve rather than via sympathetic nerves (4). At rest, these vagal HRV measures are highly correlated with respiratory sinus arrhythmia, or the degree to which heart rhythms synchronize with breathing. When inhaling, heart rate typically speeds up, and when exhaling, heart rate typically slows down, due to signals transmitted between the brain and the heart via the vagus nerve. Thus, the variability associated with better emotional well-beng is not just random noise but instead reflects heart rate oscillations synchronized with breathing.

Why should having a heart rate that responds more to breathing be associated with better emotional well-being? One potential explanation is that many of the same brain regions are involved in coordinating heart rhythms and in regulating emotions (5). Indeed, individual differences in vagal HRV have been linked with brain structures and circuits associated with emotion regulation (5-8). However, heart rate oscillations may go beyond signaling the functioning of regulatory brain regions. They may increase coordination within emotion-related brain networks, improving the brain’s capacity to regulate emotion (9). Indeed, recent findings from biofeedback studies in which people increase their own heart rate oscillatory activity suggest that episodes of high amplitude heart rate oscillations reduce stress and anxiety (10). In the typical heart rate oscillation biofeedback intervention, people slowly breathe at around 10s/breath or 0.1 Hz while receiving feedback on how much their current heart rate is oscillating in response to their breathing during daily training sessions for a few weeks (11). Breathing at this pace creates especially high amplitude heart rate oscillations because 0.1 Hz is a resonance frequency for the baroreflex system, which also produces oscillations in heart rate (12).

Intriguingly, ∼0.1 Hz oscillations in heart rate and breathing are also seen during some meditative practices (13-15), including during reciting either a yoga mantra or the rosary Ave Maria (16). Varied cultural practices may have converged on this resonance breathing frequency that creates high oscillations in heart rate because of its positive impact on well-being.

Why would daily time spent in a high physiological oscillatory state increase resting-state coordination within emotion-related brain networks? First consider what occurs during the experience of emotions or feelings. At each moment, the brain receives diverse input about current body states, with the vagus nerve serving as a primary conduit of visceral information (17-19). Mapping these body states in the brain is necessary to generate feelings even when the body state is not currently present (20). That is, people can simulate body state changes in insula and somatosensory cortices, influencing current feeling states (21, 22). This system allows for top-down modulation over feelings and emotions, as prefrontal, anterior cingulate and anterior insula regions both respond to and modulate activity in brain regions mapping visceral and somatic sensations.

Cortical brain regions involved in autonomic control including the insula and ventromedial prefrontal cortex respond to increasing or decreasing heart period intervals, supporting feedback loops that control blood flow to different areas of the body, modulate heart rate, and provide rapid responses to arterial blood pressure changes. Inducing large heart rate oscillations may serve as a workout for these feedback loops, strengthening the ability of autonomic control processes to respond to changes in somatosensory inputs, which in turn should enhance the ability to modulate fluctuations in one’s own feelings. If inducing heart rate oscillations strengthens dynamic control over emotion regulation in this way, the effects should be evident during times when the system is challenged by stimuli that induce emotions. These same feedback loops likely contribute to resting-state activity in emotion-related brain regions. Thus, daily sessions spent in a high physiological oscillatory state may also increase the coordinated activity of emotion-related resting-state brain networks (9).

Our study (ClinicalTrials.gov NCT03458910; Heart Rate Variability and Emotion Regulation or “HRV-ER”) tested the hypothesis that daily biofeedback sessions stimulating heart rate oscillatory activity in baroreflex frequencies affect the function of brain networks involved in emotion regulation, even when people are not engaged in the biofeedback. We randomly assigned 106 young adult participants to receive either ‘increase-oscillations’ (Osc+) or ‘decrease-oscillations’ (Osc-) biofeedback in daily training sessions for five weeks in a 7-week study involving pre- and post-intervention assessments (see Supplementary Fig. 1 for study schedule and Supplementary Tables 1-2 and Supplementary Fig. 2 for participant information). Participants came into the lab weekly as part of small groups and the two groups received similar rationales for their training protocols (Fig. 1a-b). We also recruited an older adult cohort whose results will be reported in a subsequent paper (due to COVID, data collection for older participants was terminated early).

**Figure 1.**
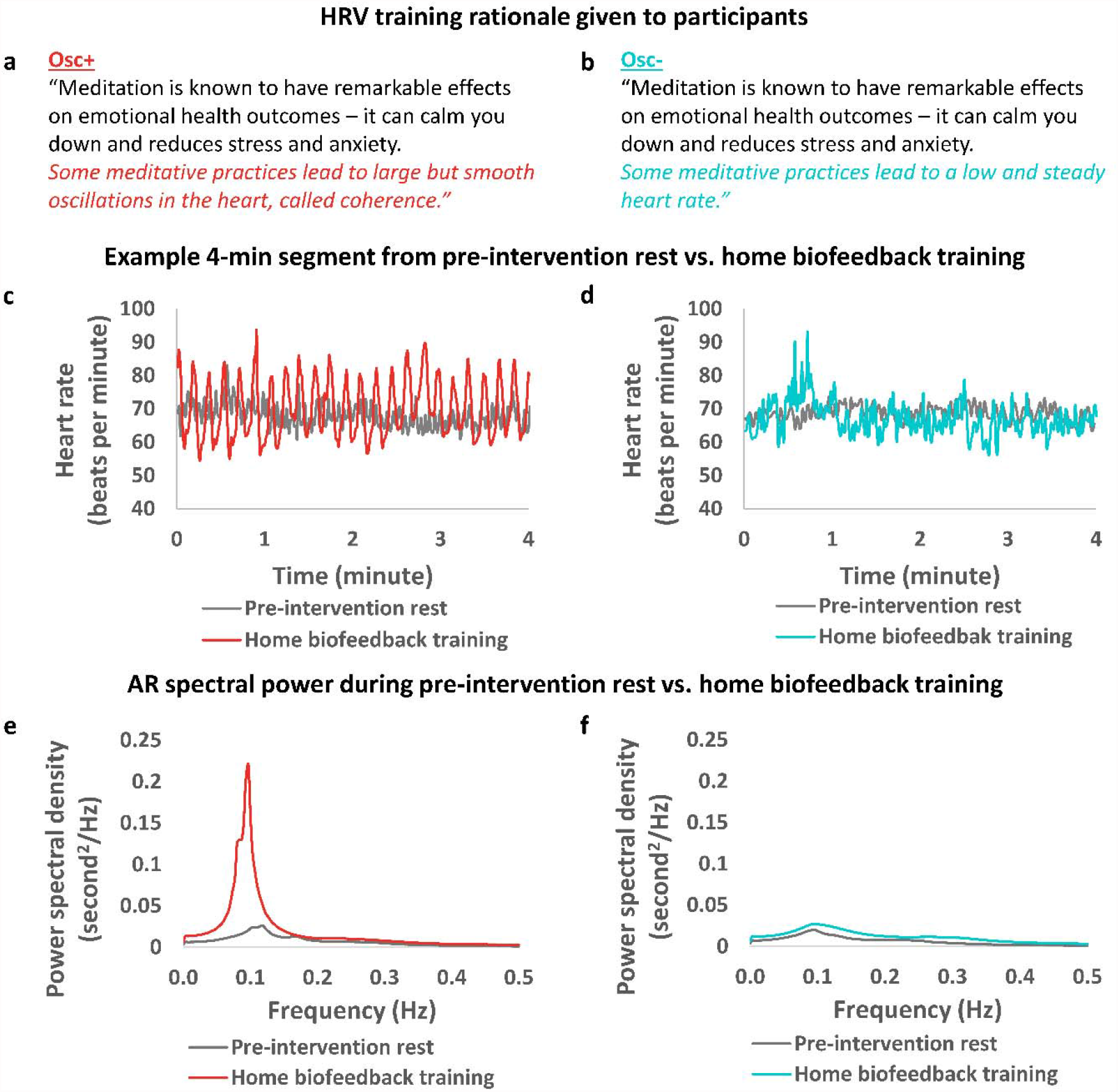
Comparisons of heart rate oscillatory activity during pre-intervention rest vs. training sessions in the two conditions. a-b: Participants received similar motivating background explanations for both conditions; c-d: Example heart rate over time during pre-intervention rest vs. home biofeedback training for an Osc+ (c) vs. an Osc- participant (d); e-f: Autoregressive (AR) spectrum shows large within-condition differences between heart rate oscillatory power during pre-intervention rest vs. home biofeedback training, for Osc+ condition (e) but not Osc- (f) condition. Note that training data in e-f was averaged across many sessions in participants’ homes (training session N = 5437), whereas pre-intervention resting-state heart rate was measured in one session per participant in the lab.

In the Osc+ condition, participants tried out several breathing paces around 10s/breath to see which induced the largest oscillations in their heart rate (their own resonance frequency; see Methods for details) (11). During their daily sessions at home, they breathed to a pacer set to this frequency while receiving biofeedback on their heart rate oscillatory activity via a ‘coherence’ score and a real-time plot of their heart rate. They returned to the lab each week to receive coaching and check again which breathing frequency produced the strongest heart rate oscillations. Assigned breathing paces ranged from 9-14s/breath.

An ideal comparison to this Osc+ intervention would be another condition with similar biofeedback information, participant expectations, and time spent training, but no increases in heart rate oscillatory activity during the training sessions. However, most relaxing states increase heart rate oscillations (23). To address this, we designed a decrease-oscillations comparison condition (Osc-) in which participants received heart rate biofeedback aimed at reducing their heart rate oscillations (summarized for them with a ‘calmness’ score and real-time feedback on their heart rate) during the training sessions. In addition, to avoid having them discover that they could reduce HRV simply by increasing physical activity (24), we asked them to also try to reduce their heart rate during the training sessions. During lab sessions, they tried out different strategies to maximize their ‘calmness’ scores during biofeedback and were advised to use their most successful strategy in their home training sessions that week. Example segments of heart rate during rest and during home training are shown for one participant from each condition (Fig. 1c-d).

A challenge for investigating how HRV biofeedback affects brain functioning is that the blood oxygen level dependent (BOLD) signal is sensitive to changes in breathing rate and carbon dioxide (CO2) levels. Thus, the main targets of our investigation were the effects of the biofeedback that carried over to the rest of the day, during normal breathing. We tested our hypotheses that the Osc+ intervention would affect both the connectivity of emotion networks during rest and these networks’ responsiveness to acute challenges by comparing post-pre resting-state connectivity in emotion-related networks as well as brain activity during an emotion regulation task.

When preregistering our outcomes, we focused on amygdala-related effects of the intervention, due to our prior findings of relationships between amygdala functional connectivity and HRV (7) and findings that the amygdala is the primary target of emotion regulation control processes (25). Our main outcome measure was pre-to-post intervention changes in resting-state right amygdala functional connectivity with a medial prefrontal cortex region associated with HRV (5). As secondary emotion-related outcomes, we examined changes in up- and down-regulation of amygdala activity and self-reported emotion regulation effectiveness during viewing emotional pictures, as well as changes in ratings of emotional well-being. Secondary outcome measures also included HRV during rest and measures of cerebral blood flow. Other secondary outcome measures (e.g., decision making, stress responsivity and cognition) will be reported elsewhere. In the current report, in addition to the amygdala-focused fMRI outcomes, we also report on the broader context of how the biofeedback affected canonical resting-state networks during rest and brain activity throughout the brain during emotion regulation.

## Results

### The Osc+ intervention increased heart rate oscillations during training but was otherwise well matched with the Osc- intervention

Participants in the Osc+ vs. Osc- conditions (*N*_*Osc+*_ = 56; *N*_*Osc-*_ = 50) did not significantly differ in the average percent of weekly assigned session time they completed (*M* = 78.32%, *SE* = 3.43 and *M* = 82.74%, *SE* = 3.74, respectively), *t*(104) = -0.87, *p* = .39, *r* = .09, in the average total amount each participant earned from lab visit payments and group and individual rewards for training adherence and quality (*M* = $293.50, *SE* = $4.69 and *M* = $293.42, *SE* = $5.99, respectively), *t*(104) = 0.01, *p* = .99, *r* = .001, in the portion of their payment rewards due to small group performance (*M* = $13.13, *SE* = $2.11 and *M* = $15.34, *SE* = $1.91, respectively; see Methods for details), *t*(104) = -0.77, *p* = .44, *r* = .08, nor in their post-intervention self-rated difficulty of training, effort, expectations, or plans to continue the intervention techniques (Supplementary Fig. 3). There was also no significant effect of condition on heart rate during home training sessions, *F*(1,95) = .73, *p* = .39, *r* = .09. However, as intended, the Osc+ participants increased their heart rate total spectral frequency power during training, *t*(51) = 9.26, *p* < .001, *r* = .54; Fig. 1e), whereas the Osc- participants did not significantly influence this metric compared to their own baseline rest (log transformed autoregressive power difference, *t*(44) = 1.39, *p* = .17, *r* = .11; Fig. 1f), leading to a significant interaction of session type (baseline vs. training) and condition, *F*(1,95) = 30.37, *p* < .001, *r* = .49. In the resonance breathing frequency range (8-16s; .063 Hz∼0.125 Hz), the two conditions showed large differences in power during training, *F*(1,95) = 43.45, *p* < .001, *r* = .56. Prior research using sympathetic versus parasympathetic blockade indicate that such increases in spectral power during slow-paced breathing are almost entirely vagally mediated (26).

### The Osc+ intervention increased functional connectivity in emotion-related resting-state networks

Quantification of functional connectivity within 18 canonical resting-state networks revealed that the two HRV biofeedback conditions also affected functional connectivity within emotion-related networks during rest. A 2 (condition: Osc+, Osc-) X 2 (network category: emotion/interoception, other; Fig. 2a) ANOVA yielded a significant interaction of condition and network category, *F*(1, 94) = 5.24, *p* = .024, *r* = .23. The Osc+ intervention increased functional connectivity within emotion-related networks significantly more than the Osc- intervention (Fig. 2b), whereas there were no significant differences between conditions for other categories of canonical resting-state networks (for breakdown of intervention effects across all 18 networks separately, see Supplementary Fig. 4).

**Figure 2.**
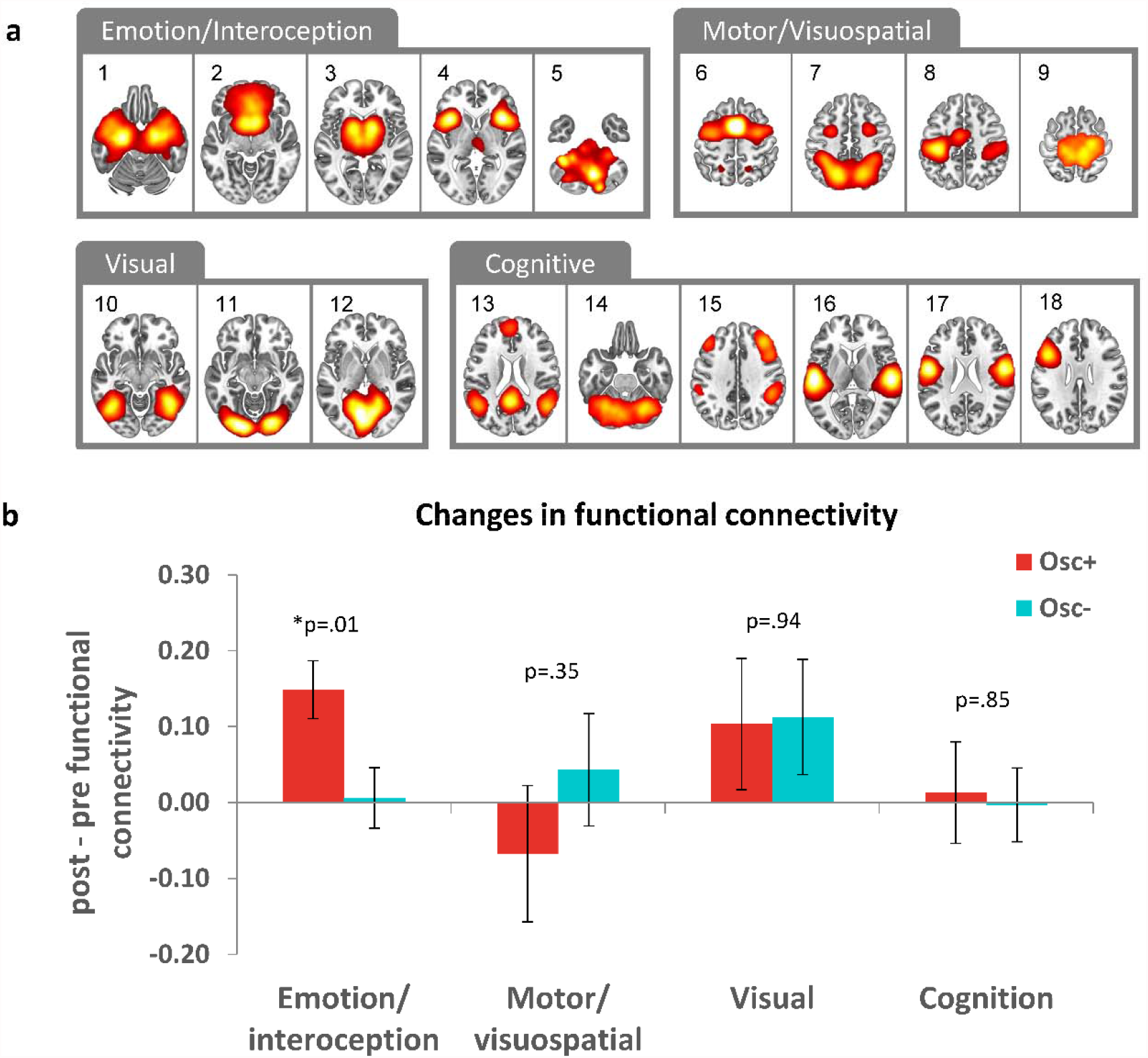
We examined changes in canonical resting-state networks (a) from pre-to post-intervention resting scans; Functional connectivity within emotion-related resting state networks also increased significantly more in the Osc+ than the Osc-condition (b). *False Discovery Rate (FDR) p<.05. Error bars indicate standard error.

Our primary outcome measure was right amygdala-medial prefrontal cortex (mPFC) functional connectivity, as this is a key emotion-related circuit (27, 28) in which functional connectivity relates to individual differences in heart rate variability (7). Seed-based analyses revealed no significant condition by time-point interaction for connectivity between mPFC and the right amygdala, *F*(1, 94) = 0.68, *p* = .41, *r* = .08 (Fig. 3a), but there was a significant interaction of condition by time-point for connectivity between mPFC and the left amygdala, *F*(1, 94) = 5.44, *p* = .02, *r* = .24 (Fig. 3b), which was driven by increased connectivity in the Osc+ condition at post intervention, *t*(48) = -2.33, *p* = .02, *r* = .26.

**Figure 3.**
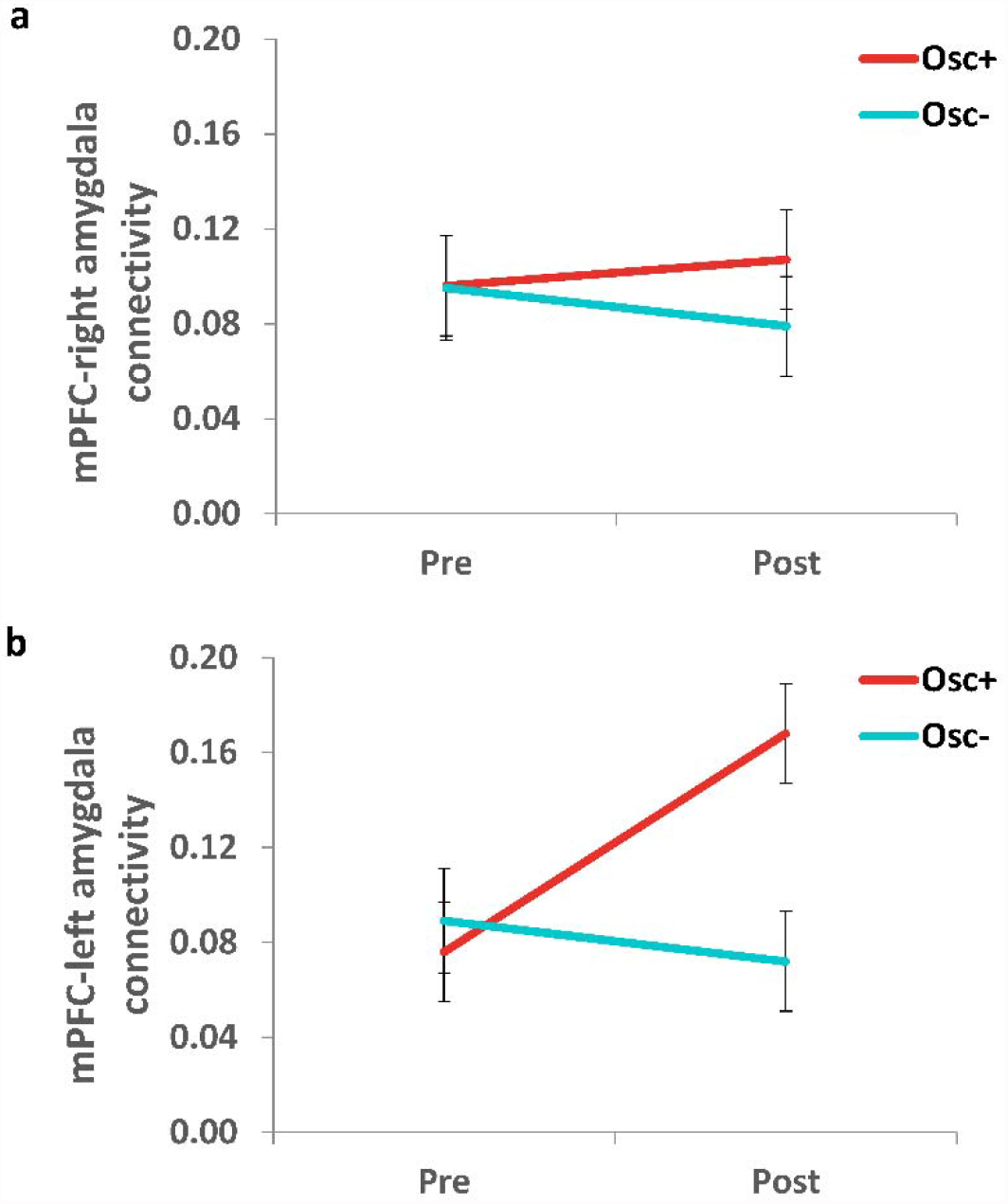
Functional connectivity between mPFC and amygdala during rest. MPFC-right amygdala functional connectivity did not differ significantly by condition (a) but mPFC-left amygdala connectivity increased during the intervention in Osc+ participants more than in Osc-participants (b).

Signal from the nearby basal vein of Rosenthal often contaminates BOLD fMRI amygdala signal (29). However, we used multi-echo imaging techniques to remove non-BOLD components such as signal from draining veins (30) and examination of our baseline whole-brain amygdala functional connectivity results indicated that our amygdala signal did not reflect signals from nearby veins (see Supplementary Figure 5).

Thus, to summarize so far, the Osc+ intervention affected functional connectivity in emotion-related brain networks, as well as functional connectivity between the left amygdala and mPFC.

### The Osc+ intervention increased down-regulation of activity in somatosensory brain regions during an emotion regulation task

Our next question was how the intervention affected the ability to regulate brain activity associated with emotional experience during externally induced emotional arousal. To test this, both before and after the intervention, participants completed an emotion regulation task during a functional scan of their brain. They saw negative, positive and neutral pictures one at a time. Before viewing each picture, they were asked to either intensify or diminish the emotional arousal the picture elicited (whether positive or negative), or to just view it (Fig. 4a). They were allowed to regulate emotions using strategies of their choice, but on post-task questionnaires over 95% of participants indicated relying on cognitive reappraisal strategies.

**Figure 4.**
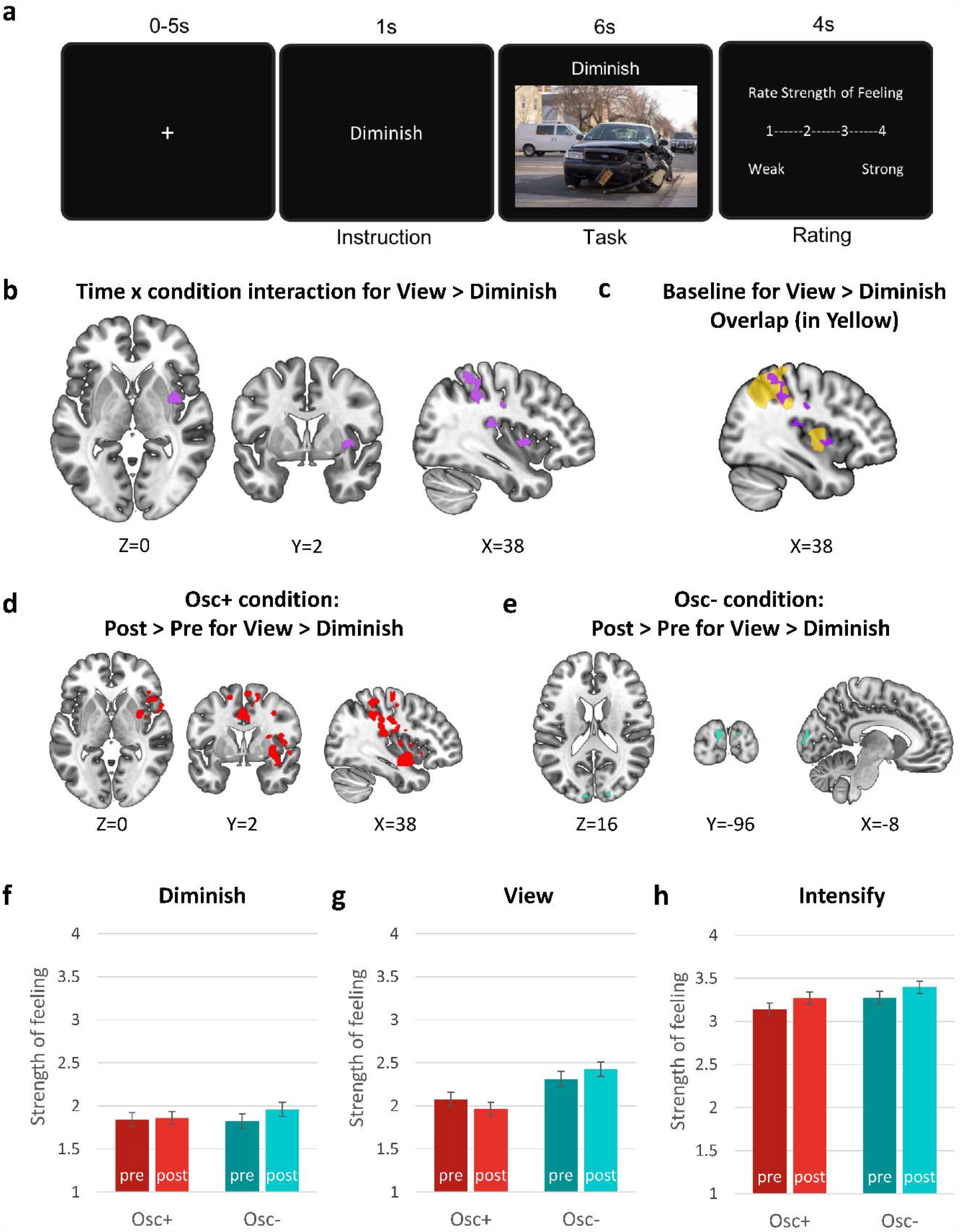
Trial design and results of the emotion regulation task. After one of the three instructions (i.e., intensify, diminish or view) was given, participants viewed each picture, performed the task, and rated the strength of feeling (a). Brain activity during Diminish trials (relative to View) showed significant time-point-by-condition interactions in somatosensory brain regions including right insula (b); brain regions that showed the intervention effect overlapped with regions that decreased activity during Diminish (relative to View) trials at baseline (pre-intervention; data from all available subjects shown for baseline in yellow, as reported in Min et al. (31)) (c); in addition, regions showing interaction effects corresponded with regions showing a decrease in activity during Diminish trials (relative to View) after the intervention in the Osc+ participants (d) but not with the occipital cluster showing a significant effect of time-point in Osc- participants (e). There were no effects of condition or time-point on ratings during Diminish trials (f); but for View trials, there was a significant interaction of condition and time-point (g). During Intensify trials, there was a main effect of time-point, with participants across conditions indicating stronger feelings in the post-than the pre-intervention scan (h).

As a manipulation check, we first confirmed that the emotional pictures affected brain activity in emotion-related regions (including the amygdala) during view trials during the pre-intervention session (see Supplementary Fig. 6 and Supplementary Table 5). In addition, based on our findings from the pre-intervention data (31) that the brain regions targeted by attempting to diminish vs. intensify emotion were mostly non-overlapping, in our whole-brain analyses, we separately compared condition effects during the Intensify trials and during the Diminish trials. We used the View condition as a baseline comparison for both.

There were no significant interactions of condition by time-point for the Intensify > View contrasts. However, for the View > Diminish comparison, there was a significant interaction of time-point and condition in clusters within the right insula, central opercular cortex, parietal operculum cortex, postcentral gyrus, supramarginal gyrus, and superior parietal lobule (Fig. 4b). These regions that showed relatively less activity during Diminish than during View at post-than at pre-intervention overlapped with the regions suppressed (relative to View) during Diminish trials at baseline across all participants (Fig. 4c; baseline results from Min et al., (31)). Comparison of post versus pre time-points for each group indicated that the time-point by condition interactions were driven by the Osc+ group who improved their ability to diminish brain activity in many interoceptive/sensory regions relative to View after the intervention (Fig. 4d). The only significant change across the five weeks in the Osc- group was in the occipital pole (Fig. 4e), but it was a cluster that did not overlap spatially with the condition-by-time-point interaction effect shown in Fig. 4b (see Supplementary Table 6 for the list of clusters). We also examined whether the intervention effect for the View > Diminish contrast differed between positive and negative emotion but did not find any significant differences.

### The two conditions had opposing effects on self-rated emotional intensity during just viewing pictures, but no differential ratings during explicit emotion regulation

Subjective ratings during the explicit regulation (diminish and intensify) trials were similar across the two intervention conditions and both groups rated pictures as more intense on Intensify trials after the intervention than before the intervention, *F*(1, 81) = 9.03, *p* = .004, *r* = .32 (Figs. 4f, h). However, when simply viewing pictures, there was an interaction of time-point and condition, *F*(1,81) = 5.65, *p* = .02, *r* = .26 (Fig. 4g). This interaction effect appeared to be due to both the Osc+ decrease, *p* = 0.10, and the Osc- increase, *p* = 0.09, in ratings of feeling strength during View trials after intervention, although the pairwise comparisons were not significant (see Supplementary Table 7 for details). Thus, although the interventions did not differentially influence conscious emotion regulation, they had differential effects on spontaneous responses to emotional pictures, potentially indicating changes in implicit emotion regulation (32). Group differences in change may have emerged during the View but not regulation trials (i.e., Intensify and Diminish trials) due to higher demand effects to give instruction-consistent responses during regulation trials than during View trials that constrained our ability to see change. Another possibility is that the effects of the intervention were stronger for implicit than for explicit emotion regulation processes (32), leading to decreased emotional responding during baseline View trials.

### The two daily biofeedback conditions affected subjective well-being similarly

Self-rated mood became less negative across the course of the intervention (Supplementary Fig. 7a), with no significant difference in change between conditions. Self-rated anxiety showed no significant changes nor condition differences (Supplementary Fig. 7b), while scores on a depression scale showed improvements across the intervention in both conditions (Supplementary Fig. 7c). Most previous studies examining the effects of heart rate variability biofeedback have relied on no-intervention controls (10); our findings highlight the importance of equating factors other than the critical physiological manipulations across conditions, as factors in the active intervention other than changes in heart rate variability may have an impact. One such factor influencing subjective ratings could be expectations. For both groups we framed the study as testing whether their biofeedback intervention would improve emotional well-being (e.g., Fig. 1a-b) and the two groups had similar expectations of improved well-being (Supplementary Fig. 3).

### Breathing and other potential physiological confounds did not differ during target MRI scans across conditions

There was not a significant time-point (Weeks 2 and 7) by condition interaction of breathing rates, heart rate, or HRV metrics during the resting-state fMRI scan, nor during the emotion-regulation task (see Supplementary Table 3 for means and statistical comparisons). In addition, during these two scans, neither exhaled carbon dioxide (CO2) levels nor the average variability in CO2 for the duration of the scan showed significant time-point by condition interactions. Likewise, a 2 (time-point: pre, post) X 2 (condition: Osc+, Osc-) ANOVA on whole-brain cerebral blood flow (CBF) during pseudo-continuous arterial spin labeling (pCASL) resting-state scans showed no significant effects.

In contrast with these lack of differences between conditions during rest and emotion regulation scans, we found significant differences in physiology in the ‘training-mimicking’ scan we conducted at the end of the session (see Supplementary Table 4). In terms of CBF, a 2 (scan type: pre-intervention rest, post-intervention training mimicking) X 2 (condition: Osc+, Osc-) ANOVA yielded a significant main effect of scan type, *F*(1,51) = 9.48, *p* = .003, *r* = .40, as CBF was lower during training mimicking (*M* = 39.33, *SE* = 1.08, *SD* = 7.88) than during rest (*M* = 42.45, *SE* = 1.14, *SD* = 8.32) across conditions. There was no significant main effect of condition, *F*(1,51) = .91, *p* = .35, *r* = .14, and the interaction of scan type and condition was not significant, *F*(1,51) = 1.30, *p* = .26, *r* = .17. In summary, the two conditions had significant effects on breathing, CO2, and HRV during training that did not carry over to the target scan sessions where we assessed emotion-related brain activity during rest and during emotion regulation.

## Discussion

Our study followed up on intriguing findings suggesting that HRV biofeedback improves well-being (9, 10, 12, 33) to test the hypothesis that experiencing daily sessions involving increased heart rate oscillation (the Osc+ condition) would affect both resting-state functional connectivity within emotion networks as well as the responsiveness to emotion regulation attempts in brain regions involved in emotional experience. The Osc+ intervention increased the amplitude of heart rate oscillation via slow paced breathing at approximately the frequency of the baroreflex, creating resonance (12). Previous findings indicate increases in heart rate oscillatory amplitude during resonance breathing are vagally mediated (26).

When planning this study, we selected changes in right amygdala-mPFC functional connectivity as our primary outcome measure because of our prior observation that right amygdala-mPFC functional connectivity was associated with HRV (7); we were interested in whether HRV plays a causal role in increasing functional connectivity within this circuit. In the current study, spending 20-40 minutes/day in a high physiological oscillatory state for a few weeks had no significant effect on right amygdala-mPFC connectivity, thus failing to confirm our main hypothesis. However, this intervention did increase left amygdala-mPFC functional connectivity. A prior meta-analysis identified the left (but not right) amygdala as showing activity related to HRV (5) and our prior study examining the relationship of how amygdala functional connectivity relates to individual differences in HRV found that, in younger adults, both left and right amygdala connectivity with ventrolateral PFC was related to HRV (7). Thus, prior studies have identified both right and left amygdala functional connectivity relationships with HRV.

In a recent review, we proposed that daily time spent stimulating physiological oscillatory activity should increase chronic levels of oscillatory activity in emotion-related resting-state brain networks (9). Indeed, our analyses examining the broader context of functional connectivity within canonical resting-state networks indicate that the functional connectivity changes seen in the left amygdala are not unique; instead, they are part of a general pattern in our study of increased functional connectivity in emotion-related networks in the Osc+ condition, an increase in functional connectivity that is greater than in non-emotion networks. As detailed in the supplementary section (see ‘*Resting State Functional Connectivity in 18 Networks’* section), these emotion-related networks are associated with a wide range of emotional and autonomic processes. In particular, the emotion networks that Osc+ participants showed most pronounced change in involve interoceptive processing. Thus, the current findings confirm our hypothesis that inducing large oscillations in heart rate leads to increased functional connectivity within brain networks that respond to interoceptive input and help shape emotions. Although self-rated emotional well-being was not differentially affected in this healthy sample, future work is needed to test whether increasing resting-state functional connectivity in emotion-related networks can benefit patients with affective disorders.

One of our secondary outcomes examined whether the intervention would influence participants’ ability to up- or down-regulate amygdala activity on demand. There were no significant effects of the intervention on amygdala activity during emotion regulation. However, when we examined whole-brain activity we found that the Osc+ intervention led to more effective down-regulation of brain regions associated with sensing body states when attempting to regulate emotional responses to pictures. Thus, the Osc+ intervention affected both resting state functional connectivity and task-related activity in brain regions associated with emotional and interoceptive processes.

These findings not only demonstrate that daily sessions involving high heart rate oscillatory activity affect brain activity in emotion-related brain regions the rest of the day, but also have implications for models of emotion regulation. There are different models of how cognitive appraisal (the strategy used by most participants in our study) affects amygdala activity. In one model, cognitive control regions (i.e., dorsolateral, ventrolateral and ventrolateral subregions of PFC and posterior parietal cortex) engage ventromedial PFC (vmPFC), which via its anatomical connectivity with the amygdala relays the control messages (25). This model guided our initial hypothesis that increased functional connectivity between mPFC and amygdala would increase Osc+ participants’ ability to regulate amygdala activity. However, in another model, prefrontal and parietal control regions affect amygdala by altering semantic and perceptual representations in lateral temporal areas when reappraising stimuli (25). Meta-analyses of emotion regulation studies support the latter model in which conscious reappraisal does not rely on vmPFC to influence amygdala activity (25, 34). Instead, the vmPFC may influence the amygdala more during implicit emotion regulation processes (7). If the vmPFC is not engaged in the reappraisal process, this could explain how the Osc+ intervention could increase amygdala-mPFC functional connectivity during rest but not enhance modulation of amygdala activity during reappraisal. Thus, our findings support the notion that mPFC has little impact on the amygdala during explicit emotion regulation.

If amygdala-mPFC functional connectivity has little impact on explicit emotion regulation, why is it so often disrupted in various disorders involving disordered emotion regulation, such as anxiety, bipolar disorder, and postraumatic stress disorder (PTSD) (35-38)? Implicit emotion regulation processes also play a key role in well-being (32). For instance, the ability to learn through experience that a conditioned stimulus is no longer associated with an unconditioned stimulus (extinction, a type of implicit emotion regulation) is impaired in PTSD and other disorders. In rodents, vagal nerve stimulation enhances extinction learning and plasticity in the amygdala-mPFC pathway (39, 40); future research should examine whether resonance paced breathing also enhances extinction and other types of implicit emotion regulation. Our findings that the Osc+ intervention decreased self-rated emotion intensity during View trials but not during Diminish trials also suggest it affects implicit rather than explicit emotion regulation processes.

While the Osc+ intervention did not affect the ability to down-regulate the amygdala during explicit emotion regulation, it did increase down-regulation of activity in brain regions associated with sensing somatic states. In a separate report (31), we compared brain activity during “intensify” and “diminish” emotion regulation trials across all participants in the pre-intervention session. We were guided by the hypothesis that regulatory control regions act like an affective dial, turning up activity in emotion-related regions when people attempt to intensify emotions while turning down activity in those same regions when people attempt to diminish emotions. This affective dial hypothesis had been implicitly assumed by emotion regulation researchers (including us) but had not been explicitly tested. To our surprise, intensifying and diminishing emotions targeted different brain regions, with diminishing emotions decreasing brain activity in interoceptive/ somatosensory brain regions and intensifying emotions increasing activity in other emotion-related regions. This dissociation is also reflected in the intervention results, as the Osc+ intervention affected brain activity during diminishing emotions but not during intensifying emotions.

In a meta-analysis, patients with mood and anxiety disorders reappraising negative stimuli showed more activity than controls in a set of brain regions that overlaps regions that Osc+ participants were better able to down-regulate after the intervention, including the right posterior insula, right inferior and superior parietal lobule, right postcentral gyrus, and right operculum (41), suggesting that being able to down regulate these regions on demand is associated with better mental health. These brain regions process signals from the body. Large oscillations in heart rate may strengthen feedback loops involving these brain regions, making these feedback loops more responsive during emotion regulation attempts and increasing participants’ ability to down-regulate activity in these brain regions that not only sense body states, but also simulate them, such as when viewing pictures of others (22). Our findings suggest that daily practice increasing heart rate oscillatory activity improved participants’ ability to diminish activity in brain regions involved in feeling emotional body states when they wanted to minimize their emotional reactions to stimuli.

Prior work comparing up-vs. down-regulating emotions has focused on the common control regions tapped by these processes and has not addressed the question of whether these regulatory control processes target activity in different emotion-related brain regions (42). The different emotion-related brain regions targeted by Diminish and Intensify conditions in our baseline data (31) and the finding that the Osc+ intervention increased the ability to down-regulate activity in the emotion-related brain regions targeted during Diminish trials but had no effect on the emotion-related brain regions targeted during Intensify trials argue against the field’s implicit “affective dial hypothesis” in which up- and down-regulation have opposing effects on the same emotion-related brain regions. Furthermore, they suggest that some interventions (like the Osc+ intervention) may be more effective for modulating down-regulation processes, whereas other interventions may be more effective for modulating up-regulation processes.

BOLD MRI signal is influenced by breathing and by CO2 levels, thus one obvious question is whether the condition differences in change in brain activity were mediated by participants in the Osc+ condition breathing more slowly even when not engaged in a training session. This does not appear to be the case, as there were no significant differences between the Osc+ and Osc- conditions in breathing rates during the resting-state or emotion regulation scans. Heart rate, HRV, end-tidal CO2, and blood flow also did not differ significantly during these scans (although LF-HRV during seated rest increased among the Osc+ participants; see Supplementary Fig. 8). In any case, the condition differences were not the result of a global change in BOLD signal, as we found that the Osc+ condition increased functional connectivity in emotion-related networks more than in other resting-state networks and the Osc+ intervention strengthened ability to down-regulate interoception-related brain activity specifically when trying to diminish emotions. In addition, our multi-echo fMRI scan processing pipeline helped avoid common confounding signal artifacts from the basal vein of Rosenthal in our amygdala connectivity analyses (see Supplementary Fig. 5).

One of the unique strengths of our study compared to most previous HRV-biofeedback studies was the active comparison group (Osc-) who completed an intervention resembling the target Osc+ intervention, but with minimal effects on HRV (see Fig. 1). We found that participants in both conditions showed significant decreases in negative mood states and in depression scores. Thus, the active comparison group was important in revealing that some aspects of the biofeedback protocol other than its effects on HRV were associated with improved emotional well-being. One possibility is that spending time every day in an awake quiet restful state yields emotional benefits regardless of whether the relaxing state increases physiological oscillatory activity. Another is that participants’ expectations (which were similarly positive in the two conditions) led to the improvements in self-reported emotional states. It is also possible that the CES-D is not the best depression scale to assess HRV biofeedback effects (33). In any case, these findings point to the importance of including active comparison groups with matched expectations in research examining the effects of behavioral interventions on well-being (43).

Across both conditions, more than half of the participants in our study were Asian. We recruited on campus; our Asian student overrepresentation may reflect ethnic differences in interest in participating in a study related to heart rate biofeedback and meditation. As Asians and European Americans differ in their ideal affect (44) and cardiovascular physiology differs between African Americans and European Americans (45), future studies should examine whether heart rate variability biofeedback effects differ by ethnicity.

In conclusion, we found that, in young healthy adults, daily sessions involving high amplitude heart rate oscillations affected emotion-related brain activity both when resting and when diminishing emotional responses. Repeated large heart-rate increases/decreases during biofeedback sessions provide a powerful physiological input that may act as a “workout” for cortical regions involved in physiological control, enhancing the brain’s capacity to respond in goal-consistent ways when later confronted with emotional stimuli.

## Methods

### Participants

We recruited 121 participants aged between 18 and 35 years via the USC Healthy Minds community subject pool, a USC online bulletin board, Facebook and flyers (see Supplementary Fig. 2 for more details and drop-out rates per condition; see the supplementary methods section for power considerations). Participants provided informed consent approved by the University of Southern California (USC) Institutional Review Board. Prospective participants were screened and excluded for major medical, neurological, or psychiatric illnesses. We excluded people who had a disorder that would impede performing the HRV biofeedback procedures (e.g., coronary artery disease, angina, cardiac pacemaker), who currently were training using a relaxation, biofeedback or breathing practice, or were on any psychoactive drugs other than antidepressants or anti-anxiety medications. We included people who were taking antidepressant or anti-anxiety medication and/or attending psychotherapy only if the treatment had been ongoing and unchanged for at least three months and no changes were anticipated. Gender, education, age and race were similar in the two conditions (Supplementary Tables 1 and 2).

### Overview of 7-week Protocol Schedule

The study protocol involved seven weekly lab visits and five weeks of home biofeedback training (Supplementary Fig. 1; see Supplementary Information for more details). Participants were provided with 11.6-inch laptops with the biofeedback software and ear sensor units for lab sessions and training at home.

### Biofeedback Training

#### Osc+ Condition

Participants wore an ear sensor to measure their pulse. The sensor cable was connected to a USB module plugged into a USB port on the training laptop computer. They viewed real-time heart rate biofeedback on the laptop screen via the emWave pro software (46) while breathing in through the nose and out through the mouth in synchrony with a visual pacer on the right side of the biofeedback display. The software provided positive feedback with a green light when participants were correctly breathing in time with the pacer. The real-time feedback was based on a ‘**coherence**’ score for participants that was calculated as peak power/(total power – peak power). Peak power was determined by finding the highest peak within the range of 0.04 – 0.26 Hz and calculating the integral of the window 0.015 Hz above and below this highest peak. Total power was computed for the 0.0033 – 0.4 Hz range (see Supplementary Information for more details). The pulse data recorded by emWave pro software and sensor unit was saved in a database file on the training laptop and also the database file was transferred to a remote server via internet connection by custom software. Interbeat interval (IBI) data was exported from the database file for HRV analysis.

#### Osc- Condition

The same biofeedback ear sensor device and emWave Pro software were used for recording heart rate data in this condition. However, we created custom software to display a different set of feedback to the Osc- participants (47). During each Osc- training session, a ‘**calmness**’ score was provided as feedback to the participants instead of the coherence score. The calmness score was calculated by multiplying the coherence score that would have been displayed in the Osc+ condition by -1 and adding 10 (an ‘anti-coherence’ score). The net result was that participants got more positive feedback (higher calmness scores) when their heart rate oscillatory activity in the 0.04 – 0.26 Hz range was low (see Supplementary Information for more details). The pulse data was recorded and saved the same way as in the Osc+ condition. IBI data was exported from the database file for HRV analysis.

### MRI Scan Session Order

In both the pre- and post-intervention MRI sessions, scans were conducted in the following order: 1) rest during blood oxygen level dependent (BOLD) fMRI; 2) rest during pseudo-continuous arterial spin labeling (pCASL); 3) emotion regulation task during fMRI; and 4) structural scan. The post-intervention session included an additional BOLD fMRI scan followed by a pCASL scan conducted after these four initial scans so as not to influence them. During these two additional training-mimicking post-intervention scans, participants engaged in their now-daily training practice (see below for details).

### MRI Scan Parameters

We employed a 3T Siemens MAGNETOM Trio scanner with a 32-channel head array coil at the USC Dana and David Dornsife Neuroimaging Center. T1-weighted 3D structural MRI brain scans were acquired pre and post intervention using a magnetization prepared rapid acquisition gradient echo (MPRAGE) sequence with TR = 2300 ms, TE = 2.26 ms, slice thickness = 1.0 mm, flip angle = 9°, field of view = 256 mm, and voxel size = 1.0 × 1.0 × 1.0 mm, with 175 volumes collected (4:44 min). Functional MRI scans during the emotion-regulation task and resting-state scans were acquired using multi-echo-planar imaging sequence with TR= 2400 mm, TE 18/35/53 ms, slice thickness = 3.0 mm, flip angle = 75°, field of view = 240 mm, voxel size = 3.0 × 3.0 × 3.0 mm. We acquired 250 volumes (10 min) for the emotion-regulation task and 175 volumes (7 min) for the resting-state scans. PCASL scans were acquired with TR = 3880, TE = 36.48, slice thickness = 3.0 mm, flip angle = 120°, field of view = 240 mm and voxel size = 2.5 × 2.5 × 3.0 mm, with 12 volumes collected (3:14 min; 1^st^ volume was an M0 image, 2^nd^ volume was a dummy image, and the remaining 10 volumes were 5 tag-control pairs) both during resting-state (pre and post) and training-mimicking (post) scans. This ASL approach provides high precision and signal-to-noise properties and has better test-retest reliability than pulsed or continuous ASL techniques (48).

### Pre- and Post-Intervention BOLD Resting-state Scan

Participants were instructed to rest, breathe as usual and look at the central white cross on the black screen.

### Pre- and Post-Intervention pCASL Resting-state Scan

To assess whether the intervention affected blood flow during rest, in both MRI sessions participants completed a second short resting-state scan. Participants were instructed to rest while breathing normally with their eyes open. To make visual inputs similar to those viewed during the training scan (for our analyses comparing rest vs. training scans), we presented red and blue circles alternately at a random rate (see *Training sessions during BOLD and pCASL*_section below). Participants were asked not to pay attention to these stimuli.

### Training-Mimicking Sessions During BOLD and pCASL

In the post-intervention scan session after the resting-state and emotion-regulation scans, participants completed their daily training without biofeedback during BOLD and pCASL scans. By this point, participants were well-trained, having each completed on average 57 training sessions at home. For the Osc+ group, a red and blue circle alternated at their resonance frequency. For example, if their resonance frequency was 12 sec, the red circle was presented for 6 sec followed by the blue circle for 6 sec. Participants were asked to breathe in with the red circle and breathe out with the blue circle. For the Osc- group, the stimuli were the same as the Osc+ group; however, the red and blue circles alternated at a random rate and participants were told not to pay attention to them.

### Emotion Regulation Task

Participants completed an emotion regulation task (49) in the MRI scanner, which lasted for about 10 min. Each trial consisted of three parts: instruction (1s), regulation (6s), and rating (4s). First, participants were given one of three instructions: “view”, “intensify,” or “diminish.” Then, during the regulation phase, they saw a positive, neutral or negative image. Finally, they were asked to rate the strength of the feeling they were experiencing on a scale ranging from 1 (very weak) to 4 (very strong).

Before the task, we instructed participants that the cue “intensify” would indicate they should escalate the emotion evoked by the subsequent image to feel the emotion more intensely. On the other hand, we instructed them that the cue “diminish” would indicate they should moderate the emotion elicited by the image in such a way that they felt calmer. We instructed them that the cue “view” meant they should simply look at the image without trying to change the emotion.

### Analyses

#### Heart Rate Oscillations During Training and Seated Rest

We used Kubios HRV Premium 3.1(50) to compute autoregressive spectral power for each training session and for the baseline rest session (a 5-min session before lab training session) in the lab in Week 2 (Fig. 1e-f; see Supplementary Information for more details).

#### Heart Rate Oscillations and Breathing Rate During fMRI Scans

Both photoplethysmogram (PPG) and breathing data were collected using Biopac MP150 Data Acquisition System using MR-compatible sensors during resting-state and emotion regulation fMRI scans in Weeks 2 and 7. The breathing belt, TSD201 transducer, converted changes in chest circumference to electric voltage signal, which were then 0.05-1Hz bandpass-filtered, amplified with 10 times of gain, sampled at 10kHz using RSP100C. During analyses using MATLAB, the respiration signal was downsampled at 1kHz and smoothed, and two iterations of peak detection were performed to obtain an average breathing rate across each scan duration. The PPG data were collected using a Nonin Medical 8600FO Pulse Oximeter at 10kHz sampling rate and downsampled at 1kHz using MATLAB. PPG data were also analyzed using Kubios HRV Premium Version 3.1 to obtain the frequency value with peak power within the high frequency range (0.15-0.4 Hz).

#### fMRI Data

##### Preprocessing

To minimize the effects of motion and non-BOLD physiological effects, we employed multi-echo sequences during our fMRI scans. Previous work indicates that BOLD T2* signal is linearly dependent on echo time, whereas non-BOLD signal is not echo-time dependent (51). Thus, multi-echo acquisitions allow uncoupling of BOLD signal from movement artifact and significantly improve accuracy of functional connectivity analyses (52), with between 2-3 times the level of reliability of typical single-echo scans (53). We implemented a denoising pipeline using independent components analysis (ICA) and echo-time dependence to distinguish BOLD fluctuations from non-BOLD artifacts including motion and physiology (54).

##### Resting State Functional Connectivity

Seed-based functional connectivity analysis: The mPFC was defined based on a previous meta-analysis of brain regions where activity correlated with HRV (5) (i.e., a sphere of 10mm around the peak voxel, x=2, y=46, z=6). The right and left amygdala were each anatomically defined using that participant’s T1 image. The segmentation of the right and left amygdala was performed using the FreeSurfer software package version 6 using the longitudinal processing scheme implemented to incorporate the subject-wise correlation of longitudinal data into the processing stream (http://surfer.nmr.mgh.harvard.edu) (55). Labels from the specific structures (left/right amygdala) were saved as two distinct binary masks in the native space. All files were visually inspected for segmentation accuracy at each time point. We used FSL FLIRT to linearly align each participant’s preprocessed data to their brain-extracted structural image and the standard MNI 2-mm brain. We applied a low-pass temporal filter 0-0.1 Hz and extracted time series from the mPFC. For each participant, a multiple regression analysis was performed in FSL FEAT with nine regressors including the mPFC time series, signal from white matter, signal from cerebrospinal and six motion parameters. The individual amygdalae were registered to the standard MNI 2-mm brain using FSL FLIRT using trilinear interpolation followed by a threshold of 0.5 and binarize operation with fslmaths in order to keep the mask a similar size. From each participant’s mPFC connectivity map, we extracted the mean beta values from the right and left amygdalae region-of-interests (ROIs) separately, which represents the strength of functional connectivity with mPFC. Lastly, we performed 2 (condition: Osc+, Osc-) × 2 (time point: pre, post) mixed ANOVAs on functional connectivity between mPFC and the left amygdala and between mPFC and the right amygdala.

Dual regression analysis: The six motion parameters and signal from white matter and cerebrospinal fluid were removed from each participant’s preprocessed data. We used FSL FLIRT to linearly align the denoised data to each participant’s brain-extracted structural image and the standard MNI 2-mm brain. A low-pass temporal filter 0-0.1 Hz was applied in order to remove high frequency fluctuation. These data were used in a FSL dual-regression analysis (56), in which we created subject-specific time series based on spatial maps for each of 18 canonical resting state networks from a prior study (57). These individual time series were used to create subject-specific spatial maps of each network. From the subject-specific z-transformed spatial maps, we extracted mean functional connectivity values for each participant within an ROI of each of the corresponding canonical network using Laird et al’s (57) network masks thresholded at 3.1 (p < 0.001). We calculated average values within each network category (emotion/interoception, motor/visuospatial, visual, and cognitive) and computed the difference between post and pre functional connectivity values.

##### Arterial Spin Labeling

Data were preprocessed using the Arterial Spin Labeling Perfusion MRI Signal Processing Toolbox (ASLtbx) (58). M0 calibration image and 10 tag-control pairs were motion corrected, co-registered to individual participants’ T1-weighted structural images, smoothed with a 6 mm full width at half maximum Gaussian kernel, and normalized to MNI template space. Preprocessing resulted in a time-series of 5 perfusion images representing the tag-control pairs, which were averaged to create a single mean whole brain perfusion image.

We conducted voxel-wise analyses of whole brain perfusion maps in SPM12 to investigate the effects of training group and time-point on cerebral blood flow with a two-way ANOVA model. We included a study-specific gray matter mask comprised of averaged gray matter segmentations across participants’ T1-weighted structural scans in all voxel-wise analyses to restrict analyses to gray matter cerebral blood flow, as ASL has lower power to detect white matter than gray matter perfusion signal (59). An absolute threshold of 0.01 ml/100g/min was applied to remove background voxels and voxels with negative values. Following model estimation, we examined interactions of group and scan type (rest pre vs. post; rest pre vs. training), and within-group pre vs. post comparisons.

##### Emotion Regulation Data

Denoised data were analyzed using FMRIB Software Library (FSL) version 6.0.3 (60). Three levels of analyses were performed: individual BOLD signal modeling, post-pre difference within each subject, and testing the difference between groups. For each individual’s pre- and post-intervention scans, a standard general linear model estimated BOLD signal during the six seconds of emotion regulation during each trial (see Fig. 4a) with seven regressors: diminish-negative, diminish-positive, intensify-negative, intensify-positive, view-negative, view-positive, and view-neutral. Instruction and rating phases were not modeled. Intensify > view and view > diminish contrasts were conducted across trials combining positive and negative images. We also examined whether the two contrasts differed between positive and negative emotions. This first-level analysis included spatial smoothing with 5-mm FWHM, motion correction (MCFLIRT) (60), and high-pass filtering with 600s cutoff. Using a 12-degree of freedom linear affine transformation, each participant’s BOLD image was registered to a T1-weighted structural image (we registered each pre-vs. post-intervention BOLD image to the T1 image obtained in the same scan session), which was then registered to the MNI-152 T1 2mm brain image. In the second-level analysis, we used FSL’s fixed effect model to estimate the post-pre difference within subjects while controlling for the mean effect. In the third-level analysis, we performed mixed-effect analyses to compare the post-pre differences in emotion regulation conditions between the two intervention groups using FSL’s *Randomise* tool with 5,000 permutations and Threshold-Free Cluster Enhancement (TFCE) multiple comparison correction (*p* < .05) (61).

To test whether the intervention changed amygdala activity during emotion regulation, we extracted amygdala BOLD activity from the results of the above general linear model using FSL’s featquery function with binary masks of the left and right amygdala (segmented through the same method used for the resting-state scan analysis and remapped to the standard MNI 2-mm brain). The extracted BOLD activity was used as the dependent variable in an independent-means *t*-test of post-pre change across conditions and in a dependent-means *t*-test comparing intensify - view vs. view - diminish differences in amygdala activity, with Benjamini-Hochberg correction.

Mixed ANOVA models were applied to test how emotion intensity ratings changed before and after intervention and how the change differed between conditions for each trial type (Diminish, View, and Intensify; 12 trials/trial type).

## Data Availability

Data supporting the findings of this study will be made publicly available at OpenNeuro.

## Data sharing plans

Upon publication, data associated with this study will be made publicly available at https://openneuro.org/datasets/ds003823.

## Acknowledgments

This study was supported by NIH R01AG057184 (PI Mather). We thank our research assistants for their help with data collection: Michelle Wong, Kathryn Cassutt, Collin Amano, Yong Zhang, Paul Choi, Heekyung Rachael Kim, Seungyeon Lee, Alexandra Haydinger, Lauren Thompson, Gabriel Shih, Divya Suri, Sophia Ling, Akanksha Jain, and Linette Bagtas.

## Contributions

K.N., J.M., and H.Y. equally contributed to the manuscript and are co-first authors. M.M. conceptualized the study, designed the study with the input from J.F.T., P.L., and C. Chang, and analyzed the data. K.N. helped design the study, directed the research team, collected and analyzed the data. J.M. and H.Y. helped with the initial setup and design of the study, collected and analyzed the data. Data collection was also performed by C. Cho with the assistance of S.L.B., P.N. and D.W. C. Cho, S.L.B. and S.D. also analyzed the data. J.F.T., P.L., C. Chang, D.A.N. and V.Z.M. provided technical assistance with data acquisition and analyses and helped interpret the results. With the supervision of S.N., T.F. developed a customized app for the Osc- training group and contributed to data management. N.M. developed a customized app for participants to track their training progress and contributed to data management. All authors contributed to manuscript preparation.

