## Supplementary for "Increasing coordination and responsivity of emotion-related brain regions with a heart rate variability biofeedback randomized trial"

**Supplementary Information**

**Supplementary Methods**

***Power Considerations***

No prior studies had examined effects of these interventions on brain function so we were unable to estimate effect sizes based on prior neuroimaging data. We elected to power our study to detect medium or larger effect sizes. Our main planned statistical comparisons were repeated-measures ANOVAs with within-between interactions. For these, a total sample size of 46 would give 90% power to detect moderate effect sizes of f = .25 with 𝛼 = .05, given an assumed correlation among the repeated measures of .5 (1). We also planned to examine within-subject change within each of the conditions. A sample size of 44 in each group would give 90% power to detect within-group change effect sizes of d = .5 in a two-tailed t-test with 𝛼 = .05 (1). Thus, we aimed for an N = 100 completion rate across the two groups to be able to accommodate potential exclusions for movement during imaging or other data quality issues. Supplementary Figure 1 details how many participants we were able to include for each category of data.

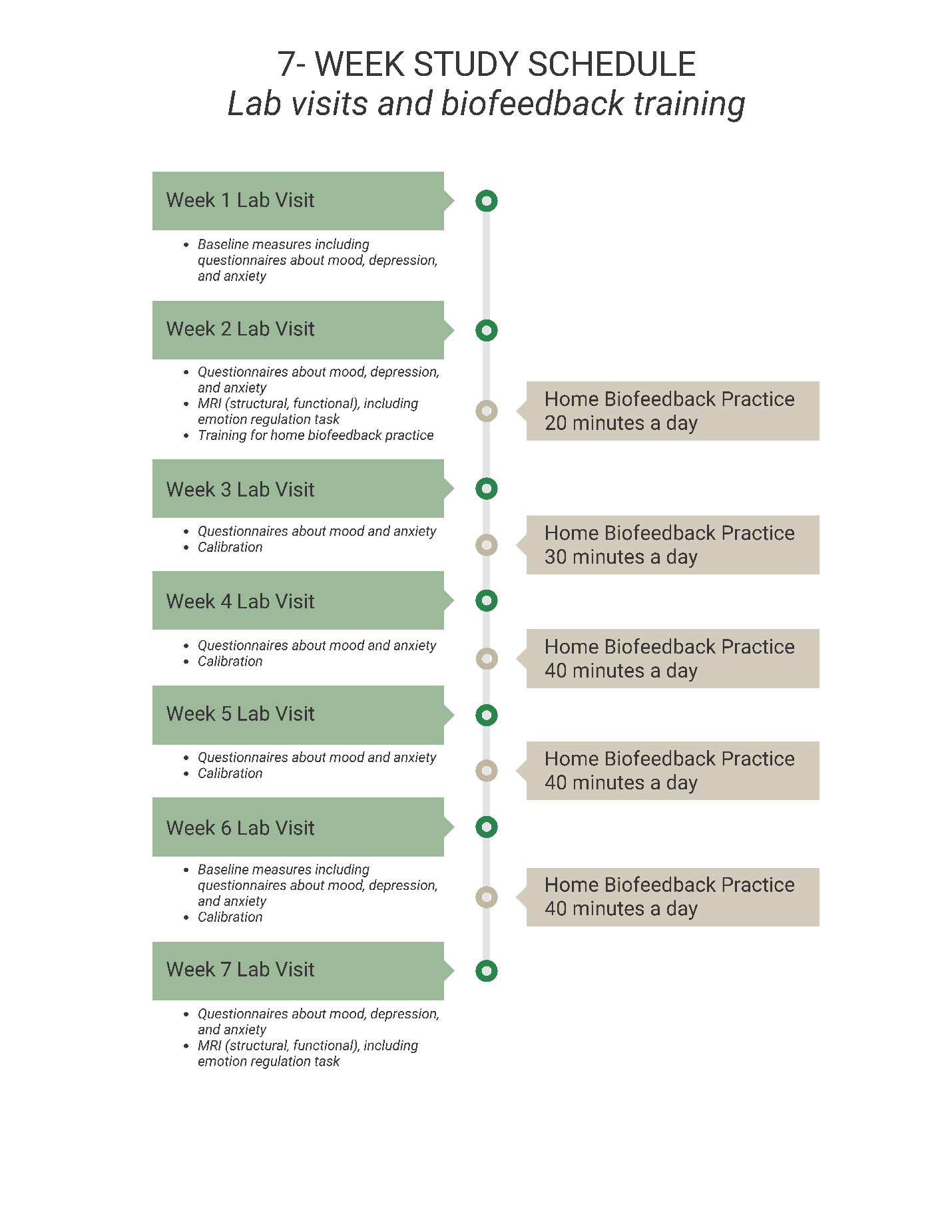

***Supplementary Figure 1. Overview of the schedule of weekly activities participants completed during the study.***

**CONSORT 2010 Flow Diagram**

### Enrollment

Randomized (n=121)

Allocated to intervention (n=58)

- Completed allocated **Osc-** intervention (n=50)
- Did not receive allocated intervention (dropped out) (n=8)

Allocated to intervention (n=63)

- Completed allocated **Osc+** intervention (n=56)
- Did not receive allocated intervention (dropped out) (n=7)

### Allocation

MRI completed (n=48)

- Not completed due to participant expressing likelihood of dropping out (n=1)
- Not completed due to unremovable body piercing (n=1)

Non-MRI measures completed

- Questionnaire
- Depression (CES-D) (n_responses_=100)
- Emotional Intensity Rating (n=46)

### Pre-Intervention Measures

MRI completed (n=55)

- Not completed due to mild claustrophobia (n=1)

Non-MRI measures completed

- Questionnaire
- Depression (CES-D) (n_responses_=112)
- Emotional Intensity Rating (n=52)

Non-MRI measures completed

- Heart rate oscillations during training and seated rest (n_sessions_=2659)
- Excluded due to outlier removal or technical issue with device/software (n_sessions_=83)
- Questionnaires
- Mood (POMS) (n_responses_=344)
- Missing data (n_responses_=6)
- Anxiety (SAI) (n_responses_=343)
- Missing data (n_responses_=7)

Non-MRI measures completed

- Heart rate oscillations during training and seated rest (n_sessions_=2785)
- Excluded due to outlier removal (n_sessions_=307)
- Questionnaires
- Mood (POMS) (n_responses_=391)
- Missing data (n_responses_=1)
- Anxiety (SAI) (n_responses_=390)
- Missing data (n_responses_=2)

### Throughout Study

- Practice completion (n=56)
- Heart rate oscillations during training and seated rest (n=52)
- Excluded due to device failure and outlier removal (n=4)
- Change in breathing rate during resting-state scan (n=43)
- Change in breathing rate during emotion regulation scan (n=45)
- Breathing rate during training-mimicking scan (n=42)
- Change in etCO2 during resting state-scan (n=25)
- Excluded due to unavailable equipment (n=11)
- Excluded due to poor data quality (n=13)
- Change in etCO2 during emotion regulation scan (n=24)
- Excluded due to unavailable equipment (n=11)
- Excluded due to poor data quality (n=10)
- etCO2 during training-mimicking scan (n=38)
- Excluded due to unavailable equipment (n=4)
- Excluded due to poor data quality (n=3)
- Practice completion (n=50)
- Heart rate oscillations during training and seated rest (n=45)
- Excluded due to device failure and outlier removal (n=5)
- Change in breathing rate during resting-state scan (n=41)
- Change in breathing rate during emotion regulation scan (n=34)
- Breathing rate during training-mimicking scan (n=38)
- Change in etCO2 during resting state-scan (n=21)
- Excluded due to unavailable equipment (n=12)
- Excluded due to poor data quality (n=14)
- Change in etCO2 during emotion regulation scan (n=21)
- Excluded due to unavailable equipment (n=10)
- Excluded due to poor data quality (n=9)
- etCO2 during training-mimicking scan (n=35)
- Excluded due to unavailable equipment (n=3)
- Excluded due to poor data quality (n=4)

### Analysis

### Post-Intervention Measures

MRI completed (n=48)

- Not completed due to lack of pre-intervention scan from participant who had anticipated dropping out but did not (n=1)
- Not completed due to unremovable body piercing (n=1)

Non-MRI measures completed

- Questionnaire
- Depression (CES-D) (n_responses_=98)
- Missing data (n_responses_=2)
- Emotional Intensity Rating (n=46)

MRI completed (n=52)

- Not completed due to mild claustrophobia (n=1)
- Not completed due to insufficient practice time (n=3)

Non-MRI measures completed

- Questionnaire
- Depression (CES-D) (n_responses_=112)
- Emotional Intensity Rating (n=52)
- Heart rate variability spectral frequency analyses of resting-state scan (n=36)
- Excluded due to outlier removal (n=3)
- Resting state functional connectivity (n=49)
- Excluded due to unsuccessful denoising pipeline results (n=1)
- Excluded due to slow breathing (n=2)
- Arterial spin labeling (n=33)
- Excluded from PreRest vs PostRest analysis (n=16)
- Excluded for PreRest image orientation processing error (n=2)
- Excluded for PreRest signal dropout and PostRest motion artifact (n=2)
- Excluded for PreRest and PostRest signal dropout (n=4)
- Excluded for PreRest signal dropout (n=1)
- Excluded for PostRest acquisition error (n=1)
- Excluded for PostRest motion artifact (n=5)
- Excluded for PostRest signal dropout (n=1)
- Excluded from PreRest vs PostTM analysis (n=19)
- Excluded for PreRest image orientation processing error (n=1)
- Excluded for PreRest image orientation processing error and PostTM signal dropout (n=1)
- Excluded for PreRest signal dropout and PostTM motion artifact (n=2)
- Excluded for PreRest and PostTM signal dropout (n=3)
- Excluded for PreRest signal dropout (n=2)
- Excluded for PostTM acquisition error (n=1)
- Excluded for PostTM motion artifact (n=2)
- Excluded for PostTM signal dropout (n=6)
- Excluded for PostTM wrong stimulus displayed (n=1)

### Analysis (cont.)

- Heart rate variability spectral frequency analyses of resting-state scan (n=37)
- Excluded due to outlier removal (n=3)
- Resting state functional connectivity (n=47)
- Excluded due to unsuccessful denoising pipeline results (n=1)
- Arterial spin labeling (n=33)
- Excluded from PreRest vs PostRest analysis (n=11)
- Excluded for PreRest image orientation processing error and PostRest signal dropout (n=1)
- Excluded for PreRest image orientation processing error (n=2)
- Excluded for PreRest & PostRest signal dropout (n=3)
- Excluded for PostRest image orientation processing error (n=1)
- Excluded for PostRest signal dropout (n=4)
- Excluded from PreRest vs PostTM analysis (n=16)
- Excluded for PreRest image orientation processing error and PostTM acquisition error (n=1)
- Excluded for PreRest image orientation processing error and PostTM signal dropout (n=1)
- Excluded for PreRest image orientation processing error (n=1)
- Excluded for PreRest and PostTM signal dropout (n=1)
- Excluded for PreRest signal dropout (n=2)
- Excluded for no PostTM scan acquired (n=1)
- Excluded for PostTM motion artifact (n=2)
- Excluded for PostTM signal dropout (n=7)
- Emotion Regulation Scan (n=45)
- Excluded due to unsuccessful denoising pipeline results (n=3)
- Excluded due to file error (n=1)
- Excluded due to participant error (n=3)
- Emotional Intensity Rating (n=44)
- Excluded due to file error (n=1)
- Excluded due to less than 50% response from participant (n=3)
- Excluded due to device error (n=2)
- Excluded due to participant providing same answer for all ratings (n=2)
- Questionnaires
- Depression (CES-D) (n_responses_=223)
- Missing data (n_response_=1)
- Mood (POMS) (n_responses_=387)
- Missing data (n_responses_=5)
- Anxiety (SAI) (n_responses_=386)
- Missing data (n_responses_=6)
- Other Measures of Heart Rate Activity (n_sessions_=333)
- Missing data due to participant no-show for lab visit (n_sessions_=3)
- Post-Study Participant Perceptions of Interventions (n=55)
- Missing data (n=1)
- Emotion Regulation Scan (n=39)
- Excluded due to unsuccessful denoising pipeline results (n=3)
- Excluded due to file error (n=1)
- Excluded due to participant error (n=3)
- Emotional Intensity Rating (n=39)
- Excluded due to file error (n=1)
- Excluded due to participant error (n=3)
- Excluded due to device error (n=1)
- Excluded due to participant error (n=2)
- Questionnaires
- Depression (CES-D) (n_responses_=197)
- Missing data (n_responses_=3)
- Mood (POMS) (n_responses_=344)
- Missing data (n_responses_=6)
- Anxiety (SAI) (n_responses_=342)
- Missing data (n_responses_=8)
- Other Measures of Heart Rate Activity (n_sessions_=268)
- Missing data due to participant no-show for or early departure from lab visit (n_sessions_=7)
- Missing data due to technical issue with device/software (n_sessions_=25)
- Post-Study Participant Perceptions of Interventions (n=49)
- Missing data (n=1)

### Analysis (cont.)

***Supplementary Figure 2***. Numbers of participants in each intervention condition, how many participants completed each measure, and how many were included vs. excluded in each analysis.

***Participants***

Participants were assigned to small groups of 3-6 people, with each group meeting at the same time and day each week. After recruitment and scheduling of each wave of groups were complete, groups were randomized to a condition. Upon completing the study, participants were paid for their participation and received bonus payments based on their individual and group performances (incentives for training were the same across conditions; see the *Rewards for Performance* section for more details).

***Overview of 7-week Protocol Schedule***

Each lab visit began with questionnaires assessing mood and anxiety (Supplementary Fig. 1). The first lab visit involved the non-MRI baseline measurements, including questionnaires. The second lab visit involved the baseline MRI session followed by the first biofeedback training session. Each of the lab biofeedback training sessions started with a 5-min baseline rest period. The weekly lab visits (except for weeks with MRI sessions) were run in small groups in which participants shared their experiences and tips about biofeedback training with other participants from the same condition, while 1-2 researchers facilitated the discussion. Outside the lab, participants used a customized social app to communicate with other members of their group and researchers about their progress on daily biofeedback training. The Week 6 lab visit repeated the assessments from the first lab visit. The final (7^th^) lab visit first repeated the baseline MRI session scans in the same order. Then, additional training-session scans were collected at the end of the scan protocol. Finally, after the scan, participants completed a post-study questionnaire.

***Biofeedback Training***

**Osc+ Condition.** During the second lab visit, we introduced participants to the device and had them complete five minutes of paced breathing at 6, 6.5, 5.5, 5 and finally 4.5 breaths/min (2). Next, we computed various aspects of the oscillatory dynamics for each breathing pace using Kubios HRV Premium 3.1 software (3) and assessed which one had the most of the following characteristics: highest LF power, the highest maximum LF amplitude peak on the spectral graph, highest peak-to-trough amplitude, cleanest and highest-amplitude LF peak, highest coherence score and highest RMSSD. Participants were then instructed to train at home with the pacer set to this frequency that appeared to best approximate their resonance frequency and to try to maximize their coherence scores.

During the third visit, they were asked to complete three 5-min paced breathing segments: the best condition from the last week’s visit, half breath per minute faster and half breath slower than the best condition. They were then instructed to train the following week at the pace that appeared most likely to be a resonance frequency based on the characteristics listed above. In subsequent weekly visits, during 5-min training segments, they were asked to try out abdominal breathing and inhaling through nose/exhaling through pursed lips as well as other strategies of their choice.

**Osc- Condition.** During the second lab visit, each participant was introduced to the device and feedback and was asked to come up with five strategies to lower heart rate and heart rate oscillations. The participant was asked to wear the ear sensor and view real-time heart rate biofeedback while they tried each strategy for five minutes. We analyzed the data in Kubios and identified the best strategy as the one that had the most of the following characteristics: lowest LF power, the minimum LF amplitude peak on the spectral graph, lowest peak trough amplitude, multiple and lowest-amplitude LF peak, highest calmness score and lowest RMSSD. Participants were then instructed to use this strategy to try to maximize their calmness scores in their home training sessions.

On the third visit, they were asked to select three strategies and try each out in a 5-min session. The strategy identified as best (based on the same characteristics used in the initial calibration session) was selected as the one to focus on during home training. In subsequent weekly visits, during 5-min training segments, they were again asked to try out strategies of their choice.

We wanted to avoid having participants figure out that one way to reduce their HRV and get positive feedback would be to do something like get up and do jumping jacks (physical activity typically decreases HRV (4)). Thus, we told Osc- participants to try to lower their heart rate in addition to lowering their heart rate oscillations and we intended to build into the feedback a minor point penalty when heart rate was the highest it had been in a short while. However, due to a coding error not detected until the study was over, this point adjustment did the opposite, giving a penalty when heart rate was the lowest it had been in the most recent 15 s. Specifically, every 5 s, a local maximum IBI was set based on the maximum IBI from the past 15 s. If, at that point, the participant’s current IBI was longer than this local maximum, the calmness score displayed for the next 5 s was the anti-coherence score - 2. Naturally, most of the time, current IBI was lower than the local maximum, and in those cases, the calmness score was the anti-coherence score +1. Thus, there was a penalty in their calmness score for moments when their heart rate was slower than it had been in any of the past 15 s. As reported in the results, average heart rate during biofeedback sessions did not differ significantly across conditions. Thus, this additional feedback appeared to have had little impact on heart rate, consistent with prior findings that biofeedback to increase or decrease heart rate has no significant impact (5).

***Post-Study Questionnaire***

After the Week-7 post-intervention scan, participants completed a questionnaire surveying their experience during the study. They provided self-ratings of difficulty of daily heart rate biofeedback training, level of effort to complete the training, expectations of the training impact on well-being, and likelihood of continuing the training after the study’s conclusion.

***Emotion Regulation Task***

Participants were instructed to intensify or diminish the emotional arousal the picture elicited (whether positive or negative), or to just view it. We asked participants to come up with their own methods to accomplish these emotion regulation goals. If participants had a hard time doing so during practice trials, we provided them with examples such as reinterpreting the situations in the image and adjusting the distance between the objects in the picture and themselves. We also instructed them not to generate an emotion opposite to the one they were experiencing. For example, they were not supposed to substitute a positive emotion for a negative one to moderate their emotion. After MRI scans, we had participants rate their confidence in accomplishing the four emotion regulation conditions (i.e., diminish-negative, diminish-positive, intensify-negative, intensify-positive) and report their emotion regulation strategies.

Trials from each condition were nested in groups of three within mini-blocks. A fixation cross with a jittering interval separated the same-condition events within each block such that two jittering intervals summed up to 4s. The blocks were separated by 5-s inter-block intervals, during which a fixation cross was displayed. This resulted in 14 blocks and 42 event-related trials in total. The blocks were presented in a pseudo-random manner such that no blocks with identical instruction nor blocks with same-valence images were presented consecutively. We selected six counterbalanced sets of 18 positive, 18 negative, and 6 neutral images from the International Affective Picture System (6) such that within each of the six sets, each picture valence type subset had the same average valence and arousal scores (positive images: mean valence = 7.2, mean arousal = 5.4; negative images: mean valence = 2.8, mean arousal = 5.4; neutral images: mean valence = 5.0, mean arousal = 2.8). Each participant was presented with one of these sets during the task scan before training and a different set after training.

***Weekly Questionnaires***

During each lab visit, participants completed the profile of mood states ((POMS 7)) and the state anxiety inventory ((SAI 8)). We used the 40-item version of POMS. Participants reported how much each item reflected how they felt at the moment using a scale from 1 (not at all) to 5 (extremely). Total mood disturbance was calculated by subtracting positive-item totals from negative-item totals. A constant value (i.e., 100) was added to the total mood disturbance to eliminate negative scores. Higher scores indicate greater negative affect. The SAI measures state anxiety using 20 statements. Participants indicated how they felt at the moment on a scale from 1 (not at all) to 4 (very much so). Scores range between 20 and 80 and higher scores indicate greater anxiety. We also administered the Center for Epidemiological Studies Depression Scale (CES-D (9)) in Weeks 1, 2, 6 and 7. Positive scores indicate greater symptoms of depression.

***Rewards for Performance***

In addition to receiving compensation of $15 per hour for each lab visit, participants were eligible to receive rewards based on individual and group performance. For individual performance rewards, each week participants had the opportunity to earn $2 for each instance (up to a maximum of 10) they exceeded their assigned target score (target scores were assigned each week and were the average of the top 10 scores earned from the previous week’s training sessions plus 0.3). Group performance rewards were earned when members of a participant’s group completed a minimum of 80% of their assigned biofeedback training minutes. For example, if a participant completed 100% of their training, they received an additional $3 for each group member who also completed 100% of their training. If a participant completed 80% of their training, they received an additional $2 for each group member who also completed at least 80% of their training. Rewards were calculated weekly, and participants received weekly updates on their earnings at their lab visit.

**Analyses**

***Heart Rate Oscillations During Training***

Heart rate data from ear sensors failed to save for the first four participants in the Osc- condition because of technical issues with the first version of the Osc- biofeedback software, leaving 102 participants’ data across the two conditions (5827 sessions). We averaged the autoregressive total spectral power from all training sessions for each participant. We excluded five outliers who on a box-and-whisker plot were above Q3 + 3 * the interquartile range on total power on pre-intervention rest (N=3), post-intervention rest (N=1), or average training (N=1), leaving an N of 97 (N_Osc+_ = 52; N_Osc-_ = 45; see Fig. 1e-f). In addition, we extracted the summed power within the 0.063~0.125 Hz range for each participant (corresponding with 8-16s, a range encompassing breathing paces used by Osc+ participants) to obtain a measure of resonance frequency oscillatory activity during biofeedback. Before conducting statistical analyses, we log transformed the power values.

***Heart Rate Oscillations, Breathing Rate and End-Tidal CO2 During fMRI Scans***

The central frequency of the HF component derived from autoregressive spectral analysis was used as an alternate estimate of the breathing rate (10). Among participants who had both breathing belt and PPG estimates of breathing, these two estimates were significantly correlated, *r*(52) = 0.95, *p* < 0.001, *r*(56) = 0.95, *p* < 0.001, *r*(44) = 0.97, *p*<0.001, and *r*(44) = 0.94, *p*<0.001 for the pre- and post-intervention resting-state scans and the pre- and post-intervention emotion regulation scans, respectively. Thus, for the subjects whose breathing belt respiration data were missing or not of good quality (*N* = 13 and *N* = 9 at pre- and post-intervention respectively for resting state, *N* = 10 and N = 6 at pre- and post-intervention respectively for emotion regulation, *N* = 4 for training-mimicking), we used the HF-HRV-derived estimate of their breathing rate. Breathing data with sudden signal drops without immediate recovery were categorized as poor quality data. For these poor quality cases, breathing rate was then substituted with HF-HRV-derived estimates or excluded if the estimates were not available. We also excluded breathing data we could not precisely synchronize with fMRI data due to failures of the scan start signal to record in the respiratory recordings. Breathing data was available for the analysis of breathing rate changes for 84 participants for resting state (*N_Osc+_* = 43; *N_Osc-_* = 41), 79 participants for emotion regulation (*N*_Osc+_ = 45; *N*_Osc-_ = 34), and 80 participants for training mimicking (*N*_Osc+_ = 42; *N*_Osc-_ = 38).

Exhaled carbon dioxide (CO2) levels were measured using Philips NM3 Monitor (Model 7900) with nasal cannula. The CO2 levels were fed to Biopac MP150 Data Acquisition System and sampled at 10kHz. After the CO2 data were shifted with a 9-second delay and downsampled to 1kHz, peak detection was performed at the end of each breath. A time series of detected peaks were used to calculate its mean and standard deviation of end-tidal CO2. CO2 data was available for 73 participants (*N_Osc+_* = 38; *N_Osc-_* = 35) during resting state, 64 participants (*N_Osc+_* = 34; *N_Osc-_* = 30) during emotion regulation and 80 participants (*N_Osc+_* = 41 ; *N_Osc-_* = 39) during training mimicking. Of these, we categorized CO2 data as poor quality if they showed sudden signal drops or stayed too low (these issues were mostly due to loosened cannulas). We excluded 27 participants (*N_Osc+_* = 13; *N_Osc-_* = 14) during resting state, 18 participants (*N_Osc+_* = 10; *N_Osc-_* = 9) during emotion regulation, 7 participants (*N_Osc+_* = 3; *N_Osc-_* = 4) during training mimicking. Thus, 46 participants (*N*_Osc+_ = 25; *N*_Osc-_ = 21) for resting state, 45 participants (*N*_Osc+_ = 25; *N*_Osc-_ = 20) for emotion regulation, and 73 participants (*N*_Osc+_ = 38; *N*_Osc-_ = 35) for training mimicking had CO2 data available for the analyses.

***fMRI Data***

**Resting State Functional Connectivity.** Out of 100 participants who completed the resting state scan before and after the intervention, two participants (one person from each condition) were excluded due to unsuccessful denoising pipeline results. Additionally, two Osc+ participants were excluded because during the post-intervention resting state scan they breathed slowly as if they were engaged in the Osc+ biofeedback. The remaining 96 participants were included in the resting-state functional connectivity analyses.

**Arterial Spin Labeling.** A total of 88 participants had available complete (i.e., pre- and post-intervention) pCASL data. Twenty-two participants were excluded due to errors in preprocessing or excessive motion, resulting in a total of 61 participants in subsequent pCASL analyses involving pre- and post- intervention scans, and a total of 53 participants in analyses involving pre-intervention and training scans.

**Emotion Regulation Data.** Ninety-eight participants (*N_Osc+_* = 52, *N_Osc-_* = 46) completed the emotion regulation scan before and after the intervention. Six participants (*N_Osc+_* = 3, *N_Osc-_* = 3) were excluded due to unsuccessful denoising pipeline results. Two participants (*N_Osc+_* = 1, *N_Osc-_* = 1) were excluded because task timing files were not saved correctly. Six participants (*N_Osc+_* = 3, *N_Osc-_* = 3) were excluded as they failed to respond to 50% or more of the trials. Eighty-four participants remained for fMRI analysis (*N_Osc+_* = 45, *N_Osc-_* = 39). For the emotional intensity rating analysis, we excluded an additional three subjects (*N_Osc+_* = 2, *N_Osc-_* = 1) whose data were collected with a malfunctioning response button device and four participants (*N_Osc+_* = 2, *N_Osc-_* = 2) who answered with the same number for all the trials. However, we included the six participants (*N_Osc+_* = 3, *N_Osc-_* = 3) for whom the fMRI denoising process was not successful. For the rating analysis, we analyzed 83 participants’ responses (*N_Osc+_* = 44, *N_Osc-_* = 39).

***Questionnaires***

For the POMS, SAI, and CES-D, we fit a series of linear mixed effects models using the packages lme4 (11) and lmerTest (12) in R Version 3.6.2 (13). For each measure, we tested fixed effects of time-point, training condition, and their interaction. For random effects, we included a random intercept for each subject, which fit the data better for all measures than did random effects structures with intercepts at the subgroup level or for subjects nested within subgroups, as determined using likelihood ratio tests. Random effects structures including random slopes led either to unidentifiable models or singular model fits. All models were fitted using maximum likelihood. Significance of fixed effects was determined using *F* tests with Satterthwaite’s approximation for degrees of freedom. For each measure, we performed post hoc comparisons of estimated marginal means of scores from week 1 with those from each successive week (2-7 for POMS and SAI; 2, 6 and 7 for CES-D) and applied a Bonferroni correction for multiple comparisons using the R package emmeans (14). All available data for all 106 participants were included for these analyses.

***Other Measures of Heart Rate Activity***

We used Kubios HRV Premium Version 3.1 to compute heart rate, the standard heart rate variability measures of low frequency HRV (LF-HRV, 0.04-0.15 Hz), high frequency HRV (HF-HRV, 0.15-0.4 Hz) and root mean squared successive difference (RMSSD) for baseline and training sessions during each lab visit. We fit separate models for heart rate, RMSSD, LF power and HF power, specifying fixed effects of time-point, training condition, and their interaction. Only data from weeks 2 and 7 were included in the statistical models to examine how each measure changed from pre- to post-training. Otherwise, we followed the same linear effect modeling set-up for these analyses as for the questionnaire data (see *Questionnaires* section above), using all available data for all 106 participants.

**Supplementary Results**

***Post-Study Participant Perceptions of Interventions***

Data for 104 participants (*N*_Osc+_ = 55; *N*_Osc-_ = 49) were included. Two participants’ responses to the questionnaire were not recorded. We conducted independent t-tests to determine if there were any differences in participants’ self-reported experience during the study based on their assigned condition. There were no significant differences between conditions. Specifically, there were no group differences regarding difficulty found with the biofeedback training, *t*(102) = -.478, *p* = .633, effort put into completing the training, *t*(102), = -1.083, *p* = .282, how pleasant the training was, *t*(102) = -1.106, *p* = .271, how much it was believed the training would impact well-being, *t*(102) = .854, *p* = .395, how much it was thought the researchers believed the training would impact well-being, *t*(102) = 1.756, *p* = .082, and plans to continue the training after the study ended, *t*(102) = .840, *p* = .403.

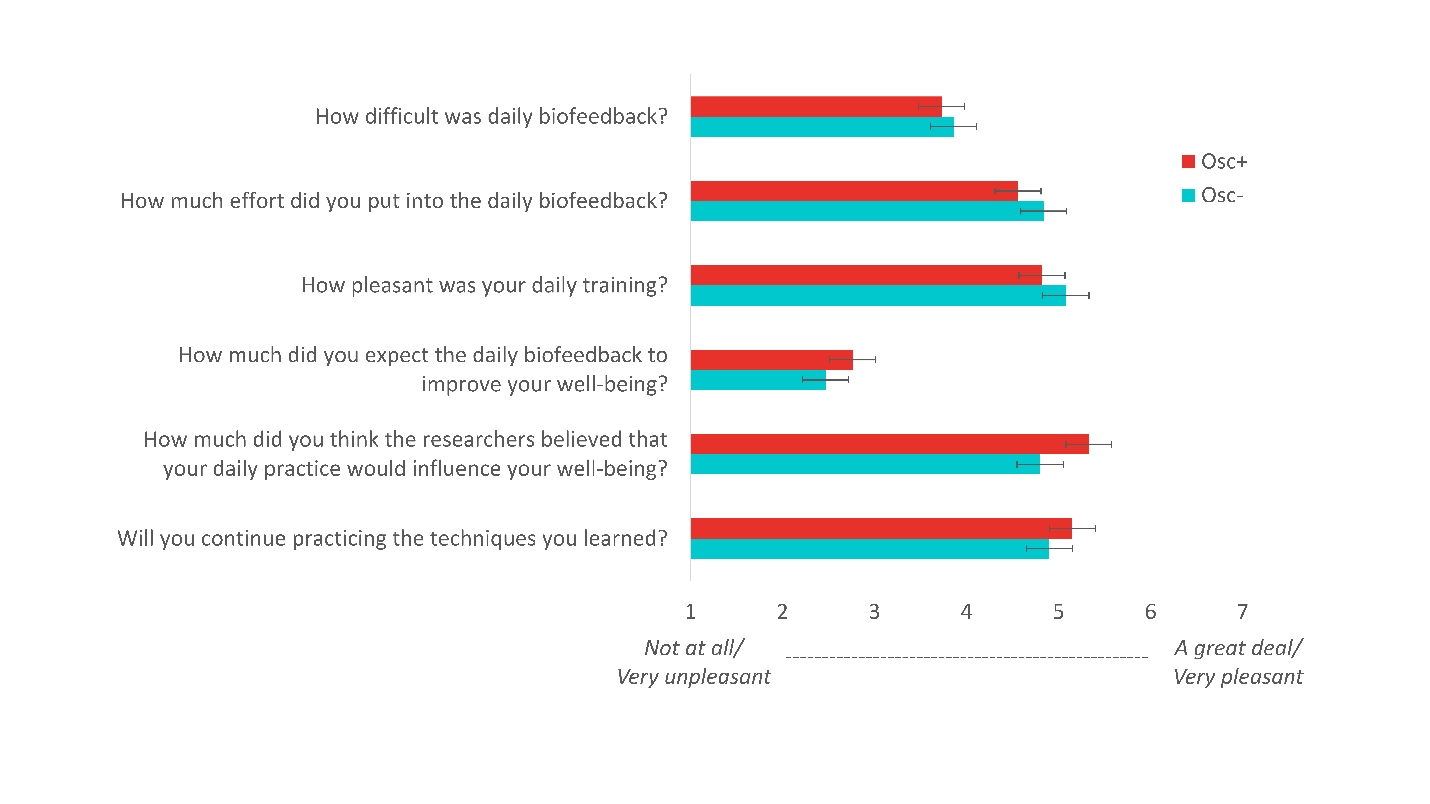

***Supplementary Figure 3***. Participant average ratings in response to post-intervention questions about their expectations and effort during the study did not significantly differ between the two conditions. Error bars indicate standard errors of means.

***Resting State Functional Connectivity in 18 Networks***

We also examined post-pre changes in resting state functional connectivity in each of the 18 networks (Supplementary Fig. 4). Two emotion networks (Networks 1 and 5) showed significant differences between conditions (*p* < .05 FDR uncorrected), which was driven by greater post-pre changes in the Osc+ condition. Network 1 includes primary olfactory and limbic association cortices, involving interoceptive processing and discrimination of emotional pictures and faces. Network 5 consists of midbrain, which is strongly associated with acupuncture and air-hunger tasks as well as interoceptive stimulation.

The other three emotion networks were Network 2 which encompasses the subgenual anterior cingulate cortex (ACC) and orbitofrontal cortex (OFC), which are associated with olfaction, gustation, and emotion, Network 3 which includes bilateral basal ganglia and thalamus, which are linked to a variety of mental processes, including reward processing, interoceptive functions, pain and somatosensory processing, and Network 4 which includes bilateral anterior insula/frontal opercula and the anterior aspect of the body of the cingulate gyrus, which are involved in executive function, complex language, affective and interoceptive processes. Although these three emotion/ interoception networks did not yield significant condition differences, the Osc+ participants showed a general increase in functional connectivity across the emotion/interoception category (i.e., Networks 3 and 4 also showed non-significant trends of p<.2 indicating relatively greater Osc+ than Osc- increases). Neither significant effects nor trends favoring the Osc+ condition were observed for any other non-emotion networks except for one cognitive network, Network 16 (*p* = .005, FDR uncorrected). This network includes the primary auditory cortices but also includes the posterior insula, which plays a key role in autonomic control. The right posterior insula has been characterized as the primary interoceptive mapping area in the cortex (15) and bilateral posterior insula responds to multiple modalities of interoceptive and sensory stimuli (16). Due to the posterior insula’s central role in interoception, it may be reasonable to include Network 16 in the category of emotion/interoceptive networks (which would further strengthen our results dissociating effects on emotion vs. other networks), although for our analyses we used Laird et al.’s (17) original categorization scheme. Thus, like the analyses in the main text, these exploratory analyses of the individual networks also suggest that the Osc+ intervention strengthened functional connectivity in brain networks associated with emotion/interoception.

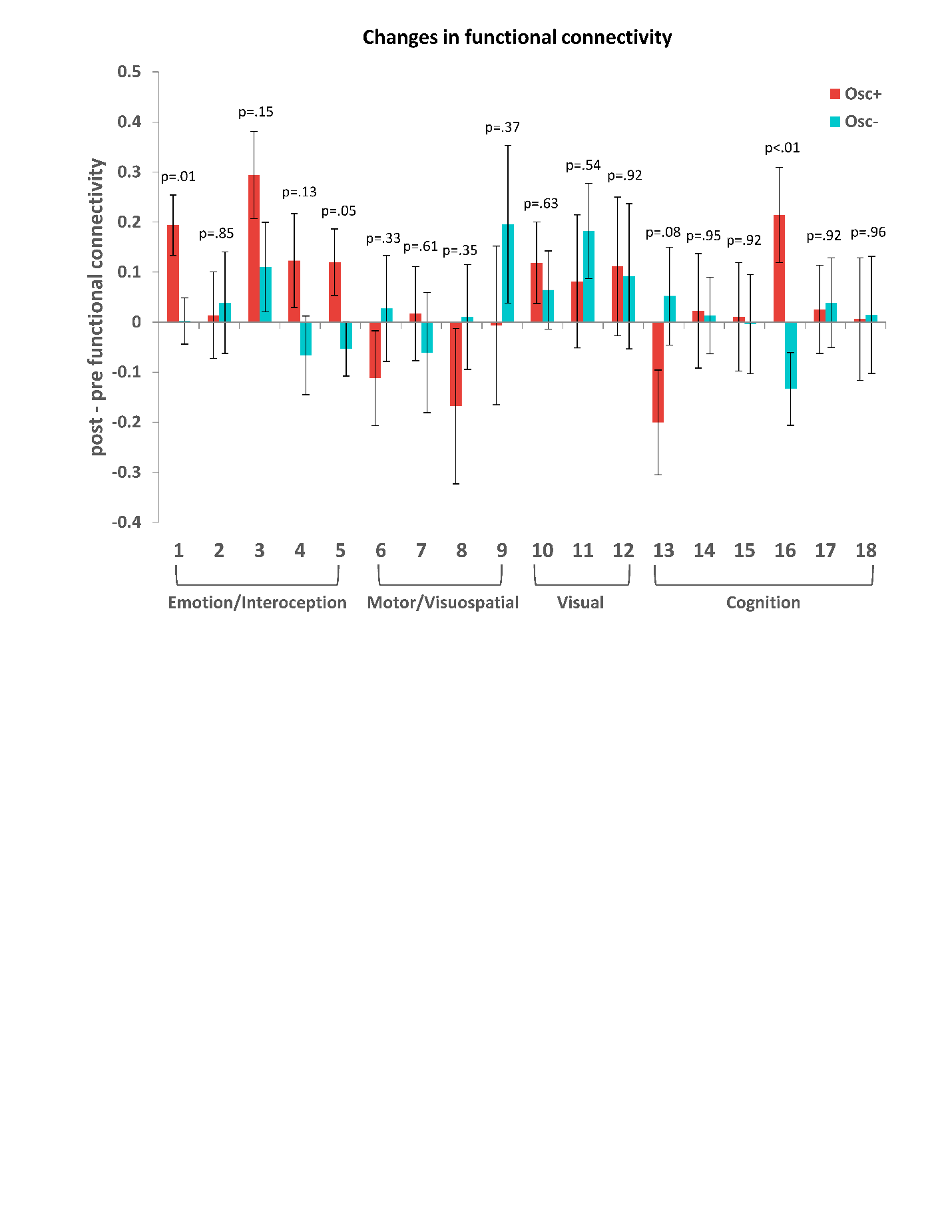

***Supplementary Figure 4.*** *Post-intervention - pre-intervention change in functional connectivity in 18 canonical resting-state networks.*

***Amygdala BOLD Signal Not Driven by Signal from Basal Vein of Rosenthal***

The multi-echo scanning sequence and its associated preprocessing pipeline optimizes the removal of non-BOLD signal, including non-BOLD cardiac artifacts (18). BOLD signal from the amygdala is often contaminated by signal from the nearby basal vein of Rosenthal (19). However, examination of whole-brain functional connectivity with bilateral amygdala ROIs in our pre-intervention data indicates that our multi-echo data collection approach and noise removal pipeline successfully avoided this confound. We used left and right amygdala as bilateral seed regions in a whole-brain functional connectivity analysis using participants’ baseline pre-intervention resting-state data and examined whether the most prominent functional connectivity was with regions overlapping the basal vein of Rosenthal, as seen in many cases (19). At the subject level, the right and left amygdala were each anatomically defined using that participant’s T1 image (for more details on the segmentation and registration, see ‘Resting State Functional Connectivity’ in the Analyses section). We applied a low-pass temporal filter 0-0.1 Hz to the preprocessed data and extracted time series from the bilateral amygdala seed regions. For each participant, we performed a multiple regression analysis in FSL FEAT with nine regressors including the bilateral amygdala time series, signal from white matter, signal from cerebrospinal and six motion parameters. The resulting images were entered into a group analysis. Due to our large baseline N yielding high power, when thresholded at Z = 3.1 (Supplementary Fig. 5a), there was widespread functional connectivity making it hard to visualize where peak connectivity was. Thus, we also display the data using a higher threshold (Z = 6) (Supplementary Fig. 5b) to facilitate comparison with the connectivity pattern to be expected if amygdala signal mainly reflects basal vein of Rosenthal signal (Supplementary Figure 5c). As expected, the highest functional connectivity signal comes from within the amygdala. In addition, the ventromedial PFC shows robust functional connectivity, validating the relevance of this region as a target in our functional connectivity analyses of intervention effects. Black arrows 1-3 indicate regions where signal would indicate contamination from veins. Unlike in the Boubela et al. (2015) figure, these regions are not among the voxels showing the strongest functional connectivity with the amygdala in our results. Furthermore, although there are a couple of clusters that appear to overlap the basal vein of Rosenthal (Supplementary Fig. 5d); these clusters center on the parahippocampal gyrus and follow its anatomical shape and so these clusters’ connectivity signal is unlikely to be driven by signal from the vein.

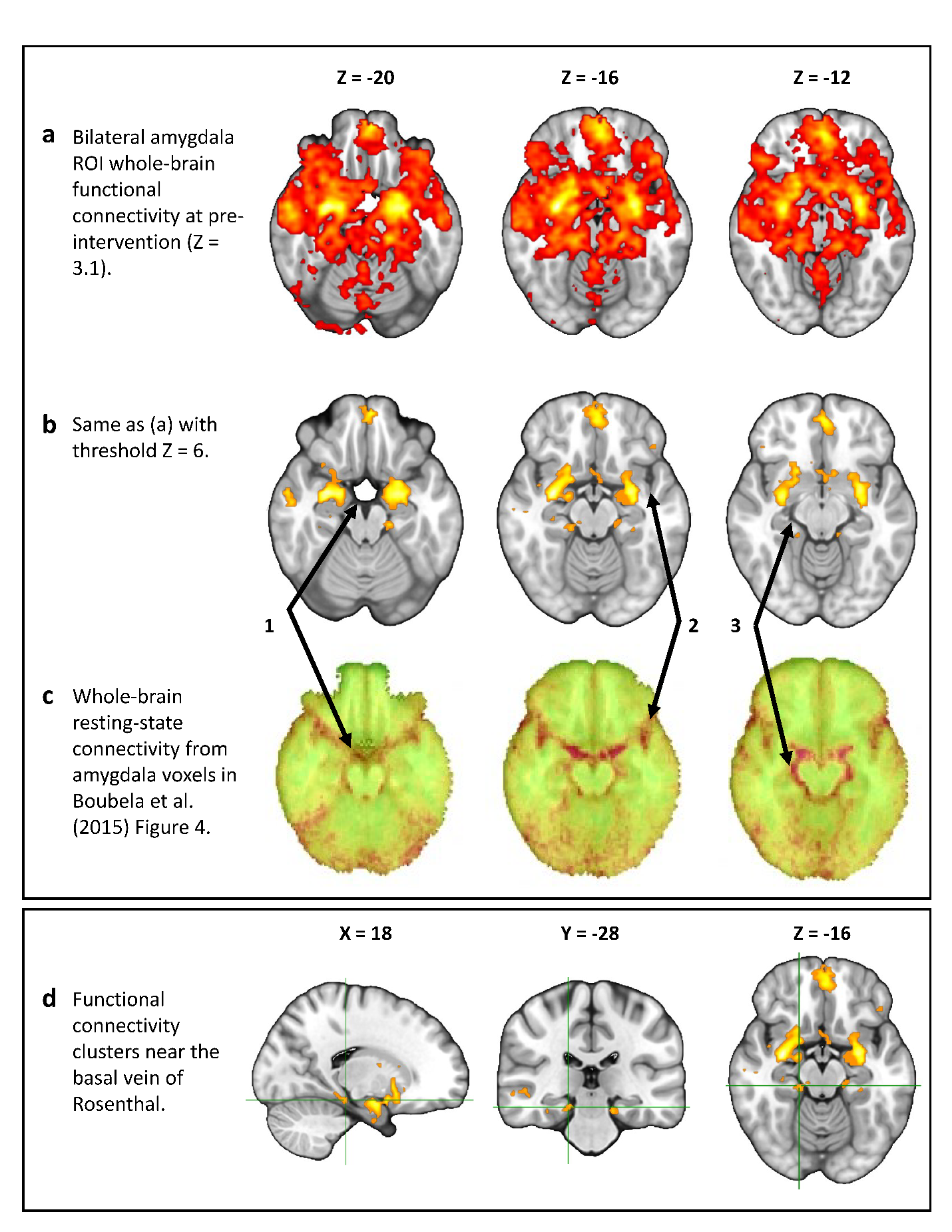

***Supplementary Figure 5*.** Whole-brain functional connectivity with bilateral amygdala at baseline during rest thresholded at Z = 3.1 (a); the same results thresholded at Z = 6 (b); and the connectivity pattern expected if amygdala signal mainly reflects basal vein of Rosenthal signal (c). Black arrow (1) indicates that dark regions corresponding with veins are not among the regions showing the strongest functional connectivity; (2) indicates that neither are the regions around the temporal pole as shown in (c); and (3) points to the edge of the brainstem at Z = -12 where Boubela et al. (19) found peak connectivity with amygdala but we did not. Row (d) provides an extended view of bilateral clusters shown in row (b) that slightly overlap with the basal vein of Rosenthal; examination of these clusters indicate that they center on the parahippocampal gyrus (see crosshairs). Panel (c) reprinted from (19).

***Emotion Regulation Task***

**Emotion Regulation Strategies Used and Confidence.** For each emotion regulation condition, more than 95% of participants used cognitive reappraisal and more than 90% of participants indicated a medium or higher confidence level.

**Impact of Emotional Pictures During View Condition.** As a manipulation check that the emotional pictures did indeed induce more emotion-related brain activity than neutral pictures, we contrasted brain activity during view trials that included emotional versus neutral pictures (regulation trials did not include neutral pictures). As shown in Supplementary Fig. 6, emotional pictures induced more activity in the amygdala as well as in other emotion-related brain regions such as the thalamus and anterior insula. As outlined in a separate report (20), we also examined differences between diminishing and intensifying emotions at baseline and found that these two processes do not target the same set of emotion-related brain regions. When participants tried to diminish emotional reactions, they were more likely to reduce activity in brain regions important for interoception whereas when they tried to intensify emotional reactions, they were more likely to increase activity in other emotion-related brain regions. Thus, despite a linear effect of down-regulation, control, and up-regulation (i.e., diminish < view < intensify) in subjective emotional intensity, a different set of emotion-related brain regions are targeted by the two regulatory processes. Given these marked baseline differences in how diminishing and intensifying emotions affect activity in emotion-related brain regions, in our fMRI analyses we examined the effects of down-regulation (i.e., view > diminish) and up-regulation (i.e., intensify > view) separately.

We first used ROI-based analyses to examine amygdala activity during emotion regulation trials. At baseline, we found significantly increased amygdala activity during intensify trials compared with view trials, *t*(83) = 3.53, *p* = 0.001, *r* = 0.39 (M = 0.10, SE = 0.02 for intensify and M = 0.04, SE = 0.02 for view) for the left amygdala and *t*(83) = 1.73, *p* = 0.09, *r* = 0.19 (M = 0.06, SE = 0.02 for intensify and M = 0.04, SE = 0.02 for view) for the right amygdala. But there were no significant differences in amygdala activity between diminish and view trials, t(83) = 0.64, p = 0.53, r = 0.06 (M = 0.03, SE = 0.02 for diminish and M = 0.04, SE = 0.02 for view) for the left amygdala and t(83) = 1.18, p = 0.24, r = 0.12 (M = 0.02, SE = 0.02 for diminish and M = 0.04, SE = 0.02 for view) for the right amygdala. Next, we examined whether post-intervention change in amygdala activity during emotion regulation trials differed between conditions. Change in amygdala activity did not show significant differences between conditions for the two contrasts, *t*(82) = 1.47, *p* = 0.15, *r* = 0.32 for view > diminish and t(82) = 0.03, *p* = 0.97, *r* = 0.01 for intensify > view in the left amygdala and *t*(82) = 0.49, *p* = 0.63, *r* = 0.11 for view > diminish and *t*(82) = -0.97, *p* = 0.34, *r* = -0.21 for intensify > view in the right amygdala (see Supplementary Table 5 for details).

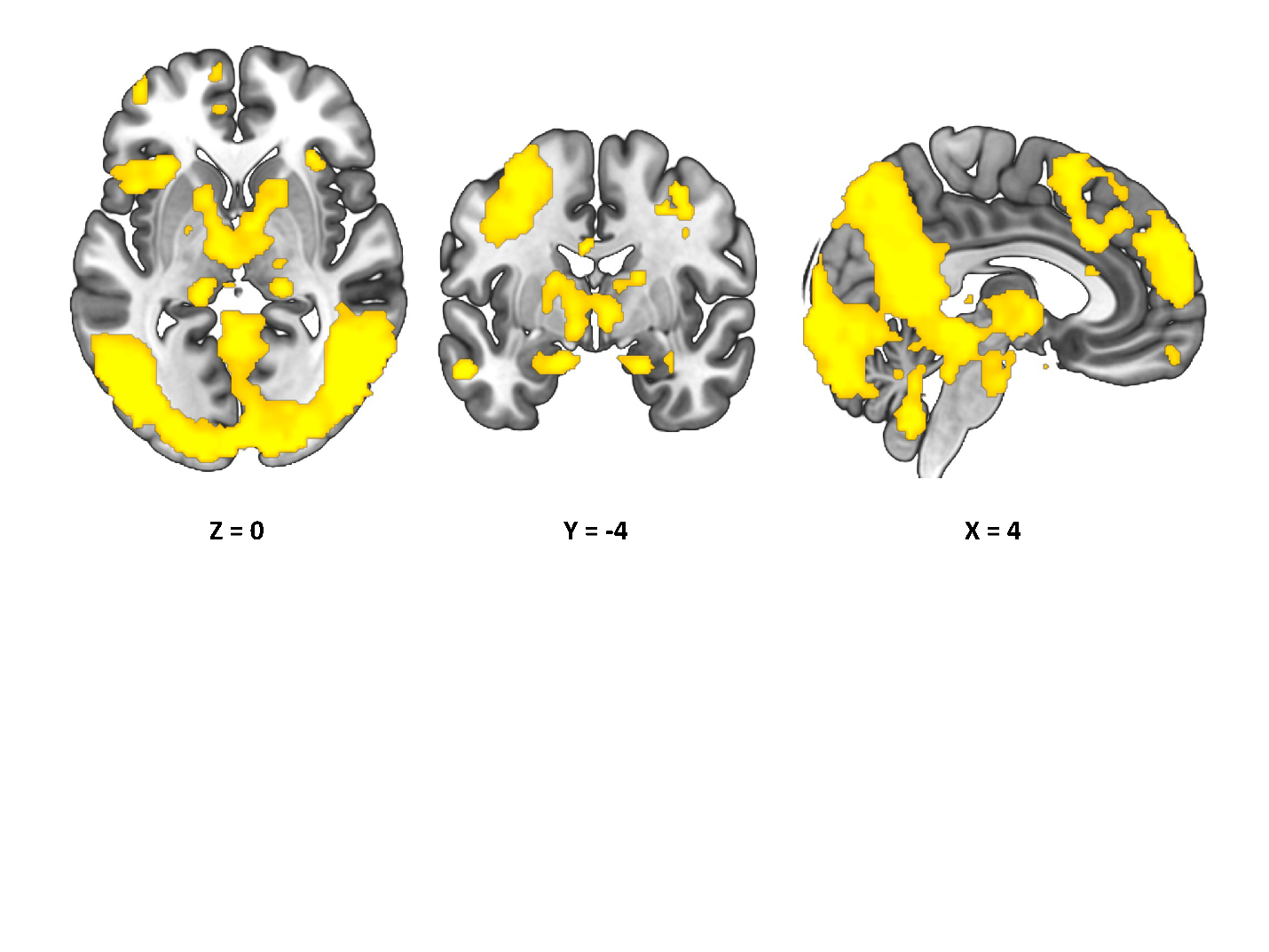

***Supplementary Figure 6*.** Brain regions significantly activated during viewing emotional vs. neutral images at pre-intervention.

***Questionnaires***

See Supplementary Fig. 7 for mean POMS, SAI, and CES-D scores across the duration of the study. Using linear mixed effects models, we found a significant main effect of time-point on POMS scores, *F*(6,625.40) = 2.47*, p* = .023, *r* = .14*,* but no significant main effect of training condition, *F(*1,106.08) = 0.05*, p* = .830*,* *r* = .02, or time-point by training condition interaction effect, *F*(6,625.40) = 0.70, *p* = .653, *r* = .08. Post hoc comparisons indicated that POMS scores were significantly lower at week 7 compared to week 1, *t*(637.46) = -3.56, *p* = .002, r = -0.14. No other post hoc comparisons yielded significant results (all *p*’s > .05). A similar approach indicated no significant effects of time-point, *F*(6,622.17) = 1.80*, p* = .097, *r* = .14, training condition, *F*(1,105.83) = 0.06*, p* = .809, *r* = .02, or their interaction, *F*(6,622.17) = 0.961, *p* = .451*,* *r* = .09, on SAI scores. Finally, we found a significant main effect of time-point on CES-D scores, *F*(3, 313.99) = 6.48*, p* < .001*,* *r* = .24, but no significant effect of training condition, *F*(1,105.72) = 0.14*, p* = .709*,* *r* = .03, or interaction effect between time-point and training condition, *F*(3,313.99) = 0.93*, p* = .425, *r* = .09. Post hoc comparisons indicated that, compared to Week 1, CES-D scores were significantly lower at Week 2, *t*(320.38) = -2.85*, p* = .014, r = -0.16, Week 6, *t*(320.38) = -2.85, *p* = .014, *r* = -0.16, and Week 7, *t*(320.38) = -4.24, *p* < .001, *r* = -0.23.

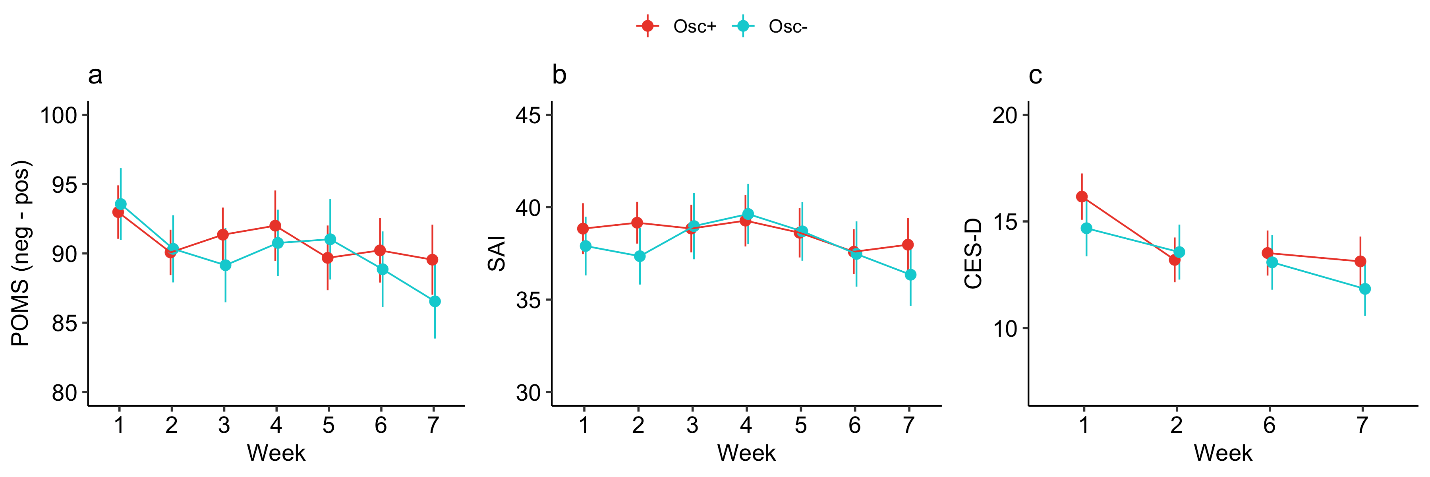

***Supplementary Figure 7.*** Mean scores on the Profile of Mood States (POMS; a), State Anxiety Inventory (SAI; b), and Center for Epidemiological Studies Depression Scale (CES-D; c) during weekly lab visits for participants in the Osc+ (red) and Osc- (blue) conditions. Higher POMS scores reflect greater negative affect, higher SAI scores reflect greater state anxiety, and higher CES-D scores reflect a higher depression quotient. A constant value of 100 was added to POMS scores to eliminate negative values. The CES-D was not administered in weeks 3, 4 or 5. Error bars reflect standard errors of the mean.

***Heart rate variability and heart rate during seated rest across study***

RMSSD, HF power, LF power and mean heart rate at seated rest across the course of the study are displayed in Supplementary Fig. 8. Using linear mixed effects models, we did not find significant main effects of time-point (pre- vs. post-intervention or Week 2 vs. Week 7), *F*(1,102) = 0.14, *p* = .714, *r* = .04, or training condition, *F*(1,102) = 1.10, *p* = .296, *r* = .10, nor a significant interaction effect between time-point and training condition on RMSSD, *F*(1,102) = 2.56, *p* = .113, *r* = .14. In contrast, this approach indicated a significant main effect of time-point on HF power, *F*(1,102) = 4.27, *p* = .041, *r* = .20, but no significant main effect of training condition, *F*(1,102) = 0.31, *p* = .576, *r* = .06, or interaction effect between time-point and training condition, *F*(1,102) = 0.54, *p* = .463, *r* = .07. In addition, we found a significant main effect of training condition, *F*(1,102) = 12.38, *p* < .001, *r* = .33, as well as a significant interaction effect between training condition and time-point, *F*(1, 102) = 4.64, *p* = .034, *r* = .20, on LF power. We did not observe a significant main effect of time-point on LF power, *F*(1,102) = 0.39, *p* = .533, *r* = .06. Post-hoc comparisons indicated significant increases in LF power from pre- to post-intervention in the Osc+ group, *t*(104) = 2.05, *p* = .043, *r* = .10, and non-significant decreases in LF power in the Osc- group, *t*(104) = -1.02, *p* = .310, *r* = .05. Finally, we found a significant main effect of time-point on heart rate, *F*(1,102) = 44.19, *p* < .001, *r* = .55^[1]^, but no main effect of training condition, *F*(1,102) = 0.01, *p* = .917, *r* = .01, or interaction effect between time-point and training condition, *F*(1,102) = 0.22, *p* = .643, *r* = .05. This pattern of results did not change when excluding participants identified as outliers based on total spectral power (see *Heart Rate Oscillations During Training* in Methods).

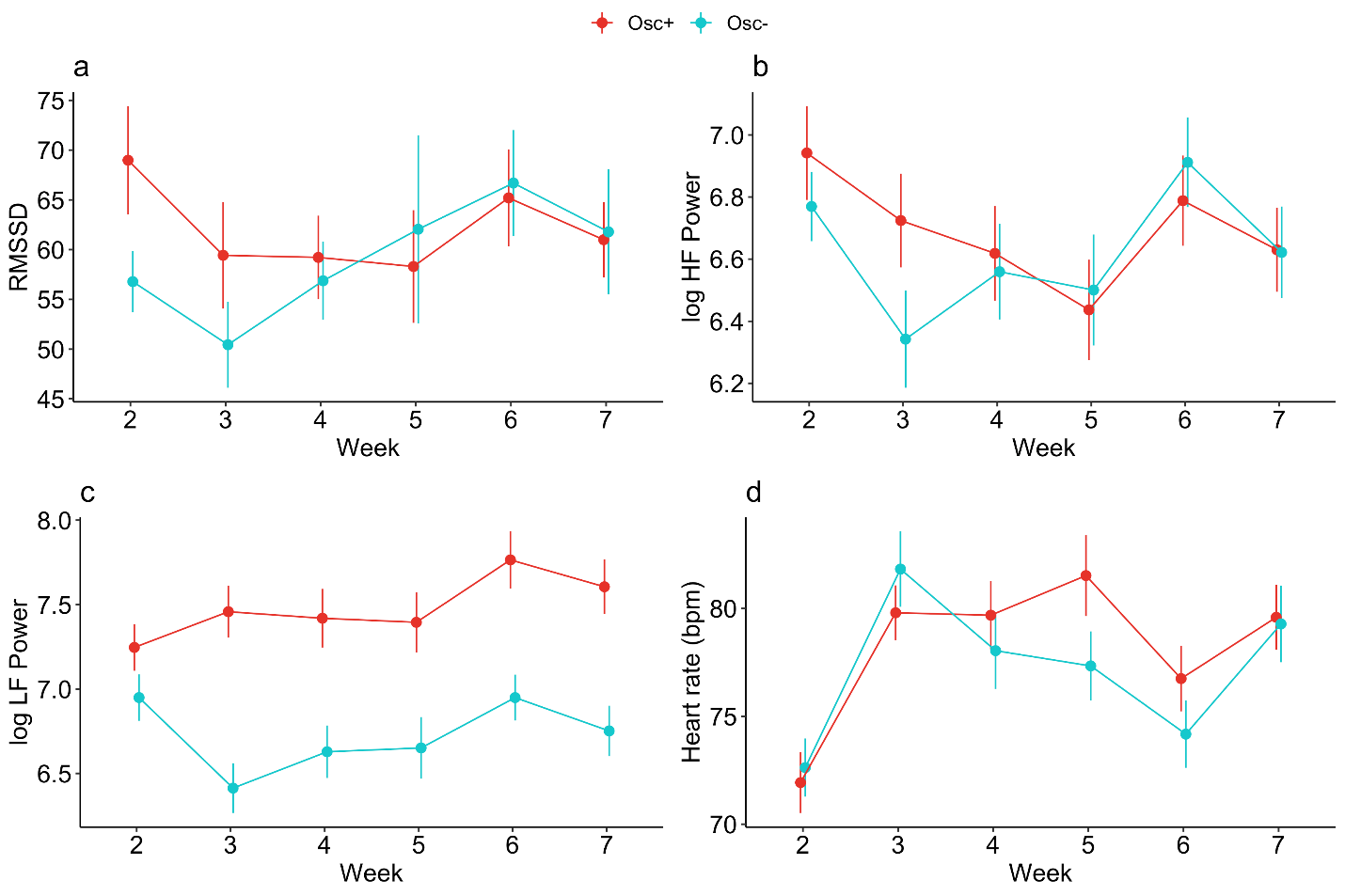

***Supplementary Figure 8***. Mean resting root mean squared successive differences (RMSSD; a), high-frequency (HF) power (b), low-frequency (LF) power (c), and heart rate (d) during weekly lab visits for participants in the Osc+ (red) and Osc- (blue) conditions. Error bars reflect standard errors of the mean.

**Supplementary Tables**

***Supplementary Table 1***. Age, sex, education, and baseline mood, anxiety and depression ratings across intervention conditions.

|  | | OSC+ (N = 56) | | OSC- (N = 50) | | | | Group difference | |
| --- | --- | --- | --- | --- | --- | --- | --- | --- | --- |
|  | | Mean (SD) | Min-Max | Mean (SD) | | | Min-Max | *p* value | |
| Age | 22.80 (2.42) | | 18-28 | | 22.60 (3.17) | 18-31 | | | .71 |
| Gender | 1.48 (0.50) | | N/A | | 1.52 (0.51) | N/A | | | .70 |
| Years Education | 16.08 (1.75) | | 12-20 | | 15.74 (2.58) | 12-24 | | | .42 |
| POMS Baseline | 92.80 (14.23) | | 68-139 | | 93.56 (18.31) | 68-166 | | | .81 |
| SAI Baseline | 38.84 (10.30) | | 21-65 | | 37.90 (11.24) | 20-68 | | | .65 |
| TAI Baseline | 42.73 (10.03) | | 22-67 | | 41.38 (12.46) | 21-70 | | | .54 |
| CESD Baseline | 15.96 (8.11) | | 4-37 | | 14.68 (9.31) | 2-45 | | | .45 |

Notes: Osc+=increase-oscillations; Osc-=decrease-oscillations; POMS=Profile of Mood States (POMS7); SAI=State Anxiety Inventory (SAI8); TAI=Trait Anxiety Inventory (TAI8); CESD=Center for Epidemiological Studies Depression Scale (9)

***Supplementary Table 2***. Race of participants across intervention conditions.

| Race | | OSC+ (N = 56) | OSC- (N = 50) |
| --- | --- | --- | --- |
| African American | 4 | | 0 |
| Asian | 41 | | 34 |
| Bi-racial | 1 | | 1 |
| Caucasian | 7 | | 12 |
| Other | 2 | | 3 |
| Prefer not to state | 1 | | 0 |

Note: among these, 2 Osc+ and 4 Osc- participants identified as being of Hispanic ethnicity.

***Supplementary Table 3***. Comparison of physiology metrics across the two conditions during pre- and post-intervention resting-state and emotion-regulation scans.

| Measure | OSC- |  | OSC+ |  | Statistical Comparison* |
| --- | --- | --- | --- | --- | --- |
|  | Pre | Post | Pre | Post |  |
| **Physiology during resting-state fMRI scan** |  |  |  |  |  |
| Breathing frequency (Hz) | .29 (.01) | .29 (.01) | .28 (.01) | .27 (.01) | *F*(1, 82) = 2.15, *p* = .15, *r* = .16 |
| End-tidal CO2 (mmHg) | 40.24 (.90) | 40.18 (.91) | 40.60 (.83) | 40.65 (.83) | *F(*1, 44) = .007, *p* = .94, *r* < .001 |
| Standard deviation of end-tidal CO2 (mmHg) | 1.37 (.16) | 1.16 (.11) | 1.47 (.15) | 1.49 (.10) | *F*(1, 44) = 0.76, *p* = .39, *r* = .13 |
| Heart rate (bpm) | 70.06 (1.47) | 68.55 (1.43) | 69.39 (1.45) | 67.18 (1.41) | *F*(1,77) = .13, *p* = .725, *r* = .04 |
| RMSSD (ms) | 45.14 (4.73) | 49.57 (6.89) | 53.47 (4.67) | 62.12 (6.81) | *F*(1,77) = .48, *p* = .49, *r* = .08 |
| LF-HRV (log of ms^2^) | 6.45 (.13) | 6.59 (.13) | 6.41 (.13) | 6.87 (.17) | *F*(1,77) = 2.0, *p* = .16, *r* = .16 |
| HF-HRV (log of ms^2^) | 6.51 (.16) | 6.59 (.17) | 6.62 (.16) | 6.87 (.17) | *F*(1,77) = .84, *p* = .36, *r* = .10 |
| **Physiology during emotion regulation fMRI scan** |  |  |  |  |  |
| Breathing frequency (Hz) | .29 (.01) | .29 (.01) | .27 (.01) | .26 (.01) | *F*(1, 77) = 1.31, *p* = .26, *r* = .13 |
| End-tidal CO2 (mmHg) | 39.13 (.90) | 39.71 (.85) | 40.25 (.84) | 39.92 (.80) | *F*(1, 43) = .54, *p* = .47, *r* = .11 |
| Standard deviation of end-tidal CO2 (mmHg) | 1.04 (.11) | 1.00 (.09) | 1.39 (.10) | 1.27 (.09) | *F*(1, 43) = .32, *p* = .58, *r* = .08 |
| Heart rate (bpm) | 72.79 (1.53) | 68.42 (1.31) | 67.94 (1.49) | 66.13 (1.27) | *F*(1, 70) = 1.57, *p* = .22, *r* = .15 |
| RMSSD (ms) | 41.77 (5.06) | 53.88 (6.79) | 61.06 (4.92) | 64.75 (6.61) | *F*(1, 70) = 1.68, *p* = .20, *r* = .15 |
| LF-HRV (log of ms^2^) | 6.44 (.14) | 6.85 (.13) | 6.65 (.14) | 6.95 (.13) | *F*(1, 70) = .44, *p* = .51, *r* = .08 |
| HF-HRV (log of ms^2^) | 6.41 (.17) | 6.74 (.17) | 6.91 (.16) | 6.93 (.17) | *F*(1, 70) = 2.09, *p* = .15, *r* = .17 |

*Notes: Statistical tests are 2 (time-point: pre, post) X 2 (condition: Osc+, Osc-) ANOVAs for scan sessions with both pre and post data. Standard errors in parentheses.

***Supplementary Table 4.*** Comparison of physiology metrics across the two conditions during the training-mimicking fMRI scan.

|  | OSC- | OSC+ | Statistical Comparison* |
| --- | --- | --- | --- |
|  | Post | Post |  |
| Breathing frequency (Hz) | .26 (.01) | .10 (.003) | *t*(78) = -17.36, *p* < .001, *r* = .89 |
| End-tidal CO2 (mmHg) | 40.74 (.79) | 39.92 (.74) | *t*(71) = -.76, *p* = .45, *r* = .09 |
| Standard deviation of end-tidal CO2 (mmHg) | 1.32 (.11) | 1.75 (.12) | *t*(71) = 2.67, *p* = .01, *r* = .30 |
| Heart rate (bpm) | 66.67 (1.35) | 65.52 (1.24) | *t*(76) = .63, *p* = .27, *r* = .07 |
| RMSSD (ms) | 59.11 (6.34) | 63.88 (5.98) | *t*(76) = -.55, *p* = .29, *r* = .06 |
| LF-HRV (log of ms^2^) | 6.93 (.13) | 8.56 (.11) | *t*(76) = -9.54, *p* < .001 , *r* = .73 |
| HF-HRV (log of ms^2^) | 6.95 (.16) | 6.45 (.19) | *t*(76) = 2.0, *p* = .03, *r* = .22 |

*Notes: Statistical tests are *t*-tests conducted to compare conditions for the training-mimicking scans which only occurred at post-intervention phase. Standard errors in parentheses.

***Supplementary Table 5*.** Activity levels within the amygdala region-of-interest (percent signal change) during the emotion regulation task.

|  |  |  | OSC+ |  |  |  | OSC- |  |  |
| --- | --- | --- | --- | --- | --- | --- | --- | --- | --- |
|  |  | Pre |  | Post |  | Pre |  | Post |  |
|  | ROI | M | SE | M | SE | M | SE | M | SE |
|  | **Left amygdala** |  |  |  |  |  |  |  |  |
|  | Diminish | 0.03 | 0.02 | 0.03 | 0.02 | 0.02 | 0.02 | 0.02 | 0.02 |
|  | View | 0.03 | 0.02 | 0.02 | 0.02 | 0.05 | 0.03 | 0.02 | 0.02 |
|  | Intensify | 0.11 | 0.03 | 0.12 | 0.03 | 0.10 | 0.03 | 0.07 | 0.03 |
|  | **Right amygdala** |  |  |  |  |  |  |  |  |
|  | Diminish | 0.02 | 0.02 | 0.08 | 0.03 | 0.02 | 0.02 | 0.02 | 0.02 |
|  | View | 0.03 | 0.02 | 0.07 | 0.02 | 0.04 | 0.02 | 0.00 | 0.03 |
|  | Intensify | 0.05 | 0.02 | 0.11 | 0.03 | 0.07 | 0.02 | 0.07 | 0.03 |

***Supplementary Table 6***. Whole-brain significant clusters and locations of local maxima during emotion regulation scan (A) time by condition interaction for View > Diminish (B) OSC+ Post > Pre for View > Diminish (C) OSC- Post > Pre for View > Diminish, with regions located based on Harvard-Oxford cortical and subcortical structural atlases.

(A) Time by condition interaction for View > Diminish

| Cluster | Voxels | *p* | *Z* | MNI (x, y, z) | | | Regions of local maxima |
| --- | --- | --- | --- | --- | --- | --- | --- |
| 4 | 1305 | 0.0386 | 3.54 | 14 | -40 | 64 | Postcentral gyrus |
|  |  |  | 3.54 | 18 | -42 | 56 | White matter adjacent to postcentral gyrus |
|  |  |  | 3.54 | 40 | -44 | 56 | Superior parietal lobule |
|  |  |  | 3.54 | 22 | -24 | 54 | White matter adjacent to precentral gyrus |
|  |  |  | 3.54 | 42 | -40 | 54 | Superior parietal lobule, posterior supramarginal gyrus |
|  |  |  | 3.54 | 14 | -42 | 54 | Precuneus cortex, postcentral gyrus |
| 3 | 126 | 0.0444 | 3.54 | -2 | -46 | 42 | Precuneus cortex, posterior cingulate gyrus |
|  |  |  | 3.54 | -2 | -52 | 58 | Precuneus cortex |
|  |  |  | 3.54 | -8 | -54 | 64 | Precuneus cortex, superior parietal lobule |
|  |  |  | 3.35 | -8 | -54 | 60 | Precuneus cortex |
|  |  |  | 3.24 | -2 | -48 | 52 | Precuneus cortex |
|  |  |  | 3.24 | -8 | -54 | 56 | Precuneus cortex |
| 2 | 65 | 0.042 | 3.54 | 34 | -2 | -2 | White matter overlapping putamen |
|  |  |  | 3.54 | 40 | -2 | 0 | Insula |
| 1 | 37 | 0.0378 | 3.54 | -18 | -60 | 56 | Superior lateral occipital cortex, superior parietal lobule |

(B) OSC+ Post > Pre for View > Diminish

| Cluster | | Voxels | | *p* | *Z* | MNI (x, y, z) | | | Regions of local maxima |
| --- | --- | --- | --- | --- | --- | --- | --- | --- | --- |
| 4 | 7245 | | 0.0026 | | 3.54 | 18 | -42 | 60 | Postcentral gyrus |
|  |  | |  | | 3.24 | 18 | -20 | 60 | White matter adjacent to precentral gyrus |
|  |  | |  | | 3.09 | 34 | -6 | -14 | White matter adjacent to amygdala |
|  |  | |  | | 3.09 | 24 | -22 | 68 | Precentral gyrus |
|  |  | |  | | 3.09 | 0 | 2 | 40 | Anterior cingulate gyrus |
|  |  | |  | | 3.09 | -18 | -56 | 58 | Superior parietal lobule |
| 3 | 78 | | 0.0424 | | 2.33 | 40 | -40 | 56 | Superior parietal lobule |
|  |  | |  | | 2.23 | 38 | -34 | 56 | Postcentral gyrus |
| 2 | 60 | | 0.0366 | | 3.04 | 46 | 28 | -4 | Frontal orbital cortex |
| 1 | 49 | | 0.0386 | | 3.04 | 64 | -14 | 16 | Postcentral gyrus, central opercular cortex |
|  |  | |  | | 2.71 | 58 | -14 | 12 | Central opercular cortex |

(C) OSC- Post > Pre for View > Diminish

| Cluster | Voxels | *p* | *Z* | MNI (x, y, z) | | | Regions of local maxima |
| --- | --- | --- | --- | --- | --- | --- | --- |
| 2 | 50 | 0.0238 | 3.54 | -8 | 100 | 6 | Occipital pole |
|  |  |  | 3.54 | -8 | -96 | 14 | Occipital pole |
| 1 | 22 | 0.0352 | 3.24 | 16 | -94 | 16 | Occipital pole |

***Supplementary Table 7*.** Ratings of feeling strength during each condition of the emotion regulation task

|  | OSC+ | | | | OSC- | | | |
| --- | --- | --- | --- | --- | --- | --- | --- | --- |
|  | Pre | | Post | | Pre | | Post | |
| Conditions | Mean | Std. Error | Mean | Std. Error | Mean | Std. Error | Mean | Std. Error |
| Diminish negative | 1.86 | 0.08 | 1.91 | 0.07 | 1.88 | 0.09 | 2.01 | 0.09 |
| View negative | 2.12 | 0.09 | 1.95 | 0.08 | 2.40 | 0.10 | 2.42 | 0.11 |
| Intensify negative | 3.15 | 0.08 | 3.35 | 0.08 | 3.26 | 0.09 | 3.41 | 0.07 |
| Diminish positive | 1.82 | 0.10 | 1.81 | 0.08 | 1.76 | 0.09 | 1.91 | 0.09 |
| View positive | 2.03 | 0.09 | 1.97 | 0.08 | 2.21 | 0.10 | 2.43 | 0.10 |
| Intensify positive | 3.14 | 0.09 | 3.19 | 0.09 | 3.29 | 0.09 | 3.38 | 0.07 |
| View neutral | 1.34 | 0.07 | 1.31 | 0.07 | 1.32 | 0.08 | 1.48 | 0.10 |

1. The increased heart rate at week 7 may be due to differences in measurement timing between Weeks 2 and 7. On Week 2, heart rate was measured approximately 30 minutes after the MRI scan since we did not want to introduce the biofeedback training, which starts with a baseline measure of heart rate, until after the scan which assessed the pre-intervention state. On week 7, most participants’ heart rate was measured within 30 minutes after their arrival to the lab (before the scan), as we did on Weeks 3-6. Heart rate was particularly low on Week 2 due perhaps to the fact that they had just been lying down in the scanner for an hour. Participants in the two conditions experienced the same protocol for order of tasks.
